# The spread of breathing air from wind instruments and singers using schlieren techniques

**DOI:** 10.1101/2021.01.06.20240903

**Authors:** Lia Becher, Amayu W. Gena, Hayder Alsaad, Bernhard Richter, Claudia Spahn, Conrad Voelker

## Abstract

In this article, the spread of breathing air when playing wind instruments and singing was investigated and visualized using two methods: (1) schlieren imaging with a schlieren mirror and (2) background-oriented schlieren (BOS). These methods visualize airflow by visualizing density gradients in transparent media. The playing of professional woodwind and brass instrument players, as well as professional classical trained singers, were investigated to estimate the spread distances of the breathing air. For a better comparison and consistent measurement series, a single high and a single low note as well as an extract of a musical piece were investigated. Additionally, anemometry was used to determine the velocity of the spreading breathing air and the extent to which it was still quantifiable. The results presented in this article show there is no airflow escaping from the instruments, which is transported farther than 1.2 *m* into the room. However, differences in the various instruments have to be considered to assess properly the spread of the breathing air. The findings discussed below help to estimate the risk of cross-infection for wind instrument players and singers and to develop efficacious safety precautions, which is essential during critical health periods such as the current COVID-19 pandemic.

## 1. Introduction

During critical periods such as the current COVID-19 pandemic, musicians have to mind not only restrictions such as social distancing and hygienic precautions, but also suffer as concerts are often not allowed. One reason is the fear that wind instruments and singers might be able to spread enormous amounts of contaminated breathing air into the room and, therefore, can be a risk for transmitting SARS-CoV-2. To assess this risk, this paper presents measurements giving an insight into how far the breathing air, which could contain infectious droplets or aerosols, spreads from different wind instruments (both woodwind and brass) and classical trained singers.

Recent studies investigated the distribution of droplets and aerosols as well as the escaping breathing air when breathing, speaking, and singing in enclosed spaces. According to the current state of knowledge, not only droplets with a size > 5 *μm* but also aerosols with a size < 5 *μm* are responsible for infections with the novel coronavirus SARS-CoV-2 (Stadnytskyi et al. 2020). While larger droplets with a size of approximately 100 *μm* fall to the ground within few seconds and being transported approximately 1.5 *m* into the room (Wei and Li 2015), smaller aerosols can remain longer in the room air and, therefore, pose a risk for room occupants. However, for singing, several studies claimed that at a distance of 0.5 *m* in front of the mouth almost no airflow can be depicted with professional singers. By placing a burning candle at 0.5 *m* in front of the singer’s mouth, it was observed that the flame hardly moved due to only small disturbances of the exhaled air (Kaehler and Hain 2020; Richter et al. 2017). Further investigations from Muerbe et al. (2020), who counted the particles emitted during professional singing, show that the emission of aerosols is much higher when singing than when speaking or breathing. For wind instruments, the exhalation of air is similar to singing: the aim is to use as little as possible breathing air to allow the vocal cords (singing), reeds (woodwind), or the lips (brass) to vibrate and stimulate the air column inside the instrument (Kaehler and Hain 2020). Here, the spreading distance is determined by the velocity of the escaping breathing air and the diameter of the outlet (Wei and Li 2015).

The present study investigates the spread of breathing air from wind instruments and singers using flow visualization and anemometry measurements. While anemometry provided the flow velocity of the escaping air, flow visualization was used to determine the spread of exhaled air into the room. Two methods of flow visualization were used: (1) schlieren imaging with a schlieren mirror and (2) background-oriented schlieren (BOS).

The optical visualization method with the schlieren mirror (in the following referred to as schlieren imaging) and the BOS method visualize density gradients due to differences in temperature or pressure in transparent media non-invasively, and therefore, with no distortion in the flow field. As the breathing air cools down while going through the instruments and spreading into the room, highly precise schlieren setups are needed.

Within building and health science, schlieren imaging has been used for the qualitative investigation of the airflow supplied by personalized ventilation (Alsaad and Voelker 2020), to investigate the propagation of the breathing air when wearing a mask (Tang et al. 2009) or during sneezing (Tang et al. 2013), or to determine the dispersion and distribution characteristics of the exhaled airflow for prediction of disease transmission (Xu et al. 2017; Tang et al. 2011). While schlieren imaging simultaneously produces high-resolution images depending on the frame rate, the measurement field is restricted to the size of the mirror (Gena et al. 2020). Since the spread of breathing air may exceed this distance, BOS was used in further investigations to avoid these spatial restrictions. BOS as a relatively young visualization method (Meier 1999) is used in numerous fields, such as the visualization of open-air explosions (Mizukaki et al. 2014), three-dimensional reconstruction of large-scale blade-tip vortices of a flying helicopter (Bauknecht et al. 2015), and instantaneous three-dimensional density field reconstruction (Nicolas et al. 2016).

The results presented in this article do not show the actual dispersion of aerosols but only the dispersion of the breathing air, which might contain infectious aerosols. Therefore, the evaluations can only be used to determine how far and to which extent the exhaled air is transported directly into the room. The evaluations shown in this article can be used as a guideline to ascertain how far the breathing air containing potentially infectious particles reaches into the room.

## 2. Visualization methods

The breathing air was visualized using the optical, non-invasive schlieren imaging with the schlieren mirror and the BOS method. Both methods allow visualizing density gradients in transparent media. These density gradients are based on differences in temperature or pressure and cause differences in the refractive index of the fluid that leads to the light rays changing their speed and, therefore, become deflected. These deflections result in discontinuous movement (Mazumdar 2013), which appears as *schlieren*, a German word meaning streaks (Settles 2001).

Since the breathing air, which escapes from the mouth of the musicians or the wind instruments, has a higher temperature or humidity (or both) than the surrounding air, the optical schlieren techniques allow visualizing these gradients. For some instruments that have a long or wide bore such as the F tuba, the air blown into the instrument cooled down when reaching the bell. Therefore, to visualize the breathing air escaping from the F tuba, the instrument was played for approximately 30 minutes to warm up the brass and consequently the air escaping the bell.

Fig. 1 shows the experimental setups of both systems. The setup of each system will be described further in the following sections.

**Fig. 1.**
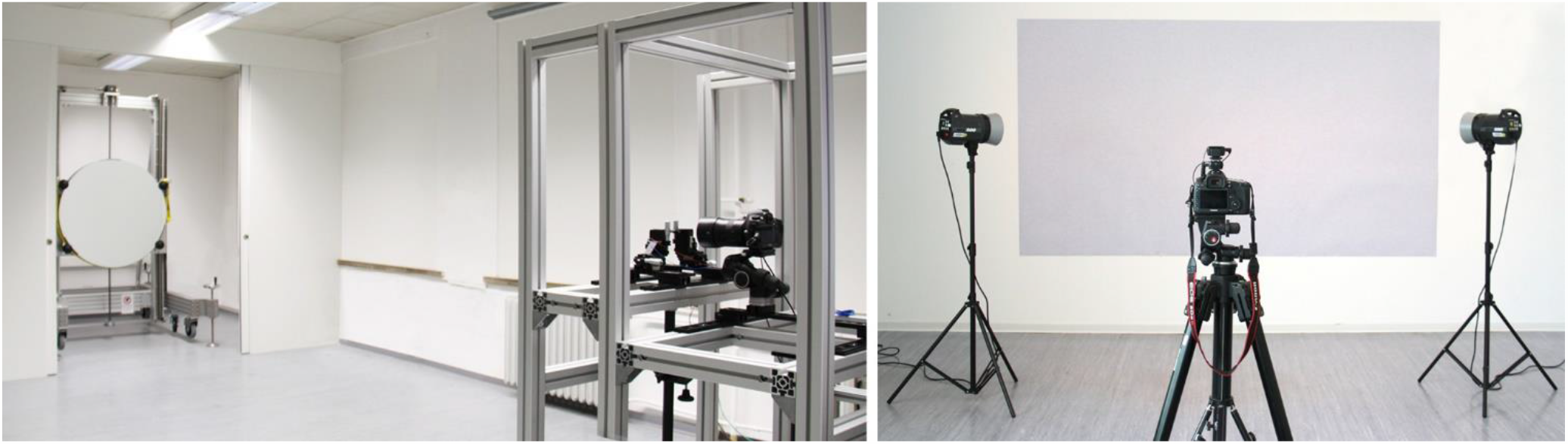
Setup of the single-mirror coincident schlieren system (left) and the BOS system (right) at the Department of Building Physics at the Bauhaus-University Weimar

### 2.1. Schlieren imaging

The single-mirror coincident schlieren system at the Department of Building Physics at the Bauhaus-University Weimar consists of a concave spherical mirror with astronomical qualities, a light-emitting diode (LED) light source, a knife-edge schlieren cutoff, and a high-resolution digital camera (Canon EOS 5DS R) (Fig. 1 left and Fig. 2).

**Fig. 2.**
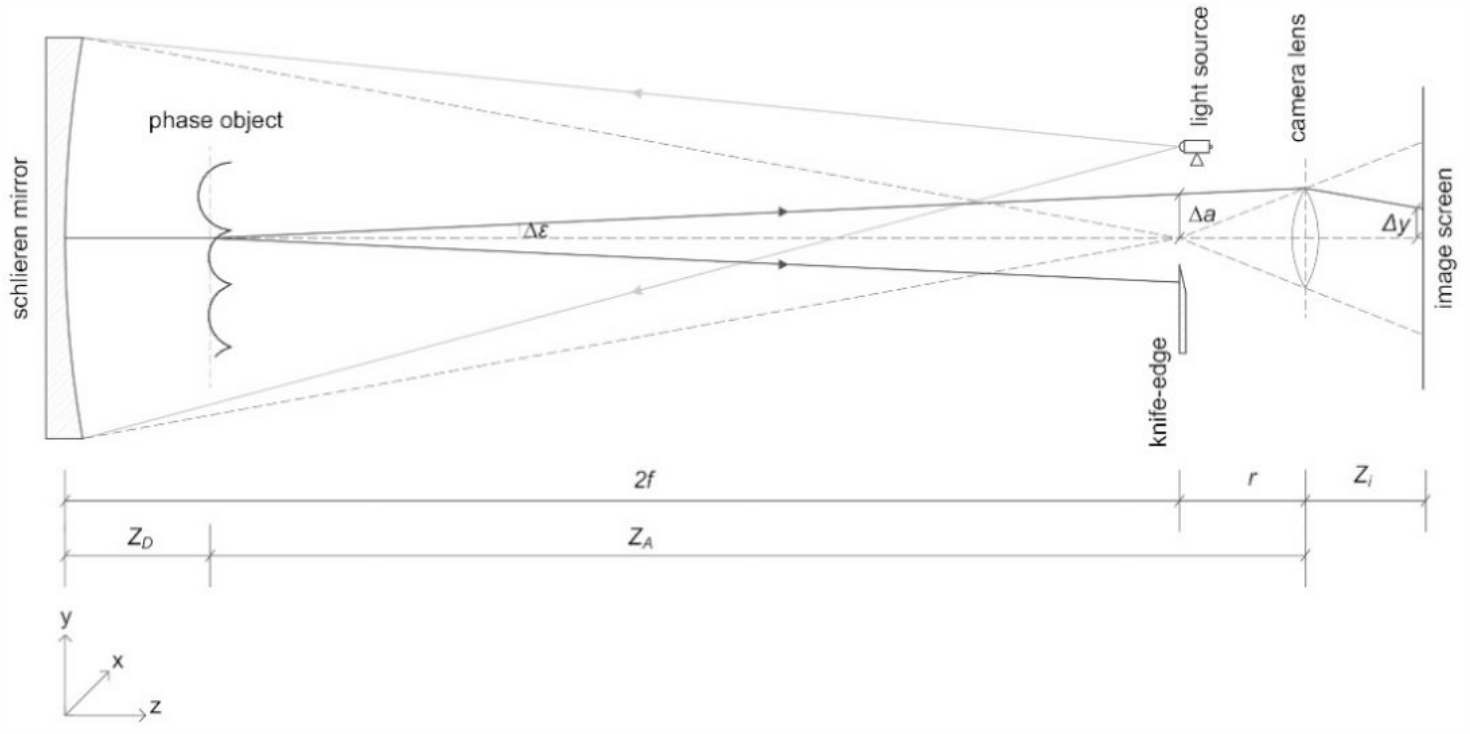
Schematic setup of the single-mirror coincident schlieren imaging system (2f ≈ 6 m)

During the investigations, the phase object (musician with instrument or singer) is placed in front of the mirror. The diverging light ray emitted from the LED reaches the mirror and returns along the coincident path where it crosses the phase object, which is inducing the density gradient. For coincident optics, the phase object is traversed twice by the same light ray. Therefore, the distance between the light source and the knife-edge was kept at a minimum distance of 5 mm to avoid double imaging. When passing through the density gradients, some rays are refracted. If these rays are refracted downwards to the extent of *Δε*, they are blocked by the knife-edge and, thus, create shadows in the image screen which constitutes the foreground of the schlieren image. On the other hand, the light rays that are refracted over the knife-edge pass to the camera sensor and brighten the image screen which constitutes the background of the schlieren image (Gena et al. 2020). In these experiments, the knife-edge was at a position where it blocks approximately 50% of the refracted light rays. Because of the low light level in the laboratory, the camera was set to a lens aperture of f/2, a shutter speed of 1/60 seconds, and ISO 320. To capture the density gradients in motion, 0.92 megapixels schlieren images (frame size 1280 × 720 pixels) were captured at 50 frames/s. This high frame rate allowed capturing still images as well as videos of the flow.

### 2.2. Background-oriented schlieren (BOS) method

The BOS system consists of a structured background (randomly distributed black squared dots on a white surface), a high-resolution camera (Canon EOS 5DS R) focusing the background, and two flashlights illuminating the background pattern (Fig. 1 right and Fig. 3). For investigations, the phase object (musician with instrument or singer) is placed in front of the structured background.

**Fig. 3.**
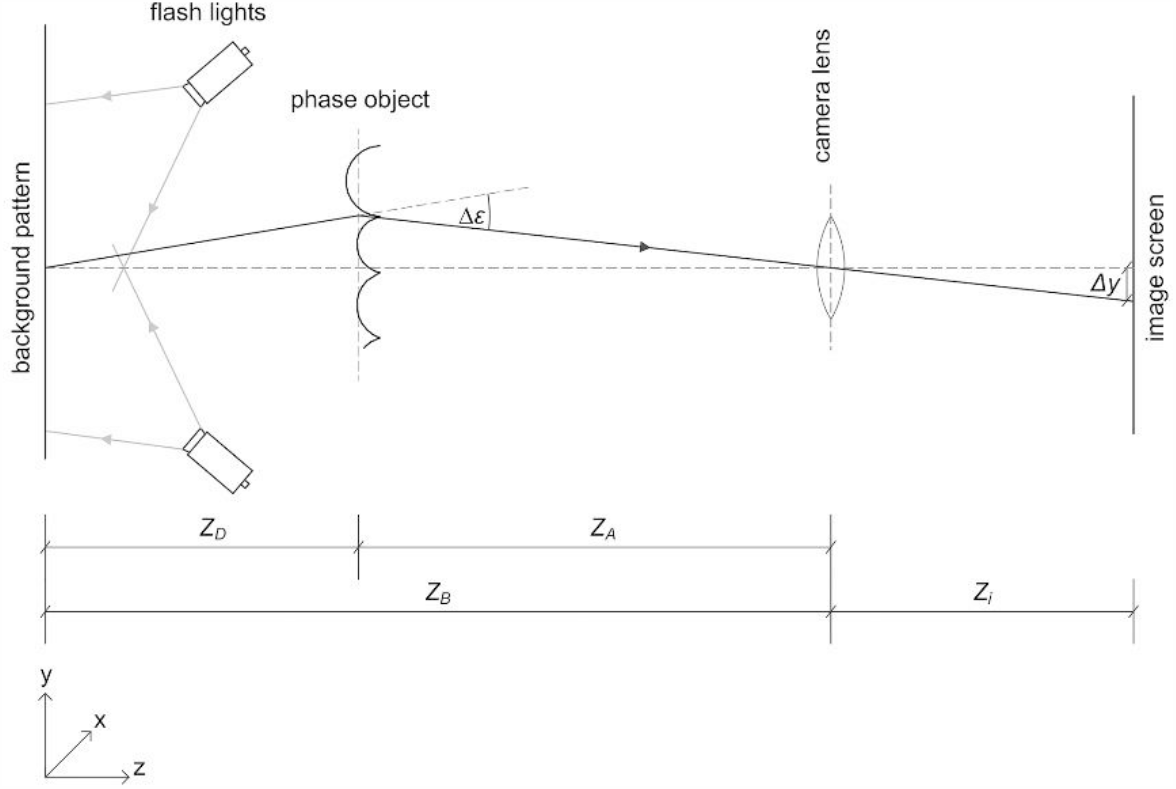
Schematic setup of the BOS system

For evaluation, two recordings of the background pattern are needed: one recording of the undisturbed background as a reference image (flow-off) and one recording of the density gradient in front of the background (flow-on). Due to the refractive index gradient, each pixel passing through the measuring field is refracted to the extent defined by the gradient. Hence, a pixel on the flow-on image appears at a different location compared to the reference image. Using cross-correlation algorithms, it is then possible to compare the intensity distributions of both images to determine the image shift of each pixel and, thus, to visualize the density gradient. Further information about the principles of BOS can be found in Becher et al. (2020).

During the investigations, the camera was set to a lens aperture of f/14, a shutter speed of 1/80 seconds, and ISO 100. To capture the density gradients, 50.6 megapixels RAW schlieren images (frame size 8,688 × 5,792 pixels) were captured at 1.5 frames/s. Due to this low frame rate, no video recordings were made with the BOS method.

### 2.3. Experimental scheme

To investigate the spread of breathing air, two separate test series were conducted. In the first series, musicians from the philharmonic orchestra *Thüringen Philharmonie Gotha – Eisenach*, Germany were positioned in front of the schlieren mirror while playing their particular instrument. Since the visualization range of the schlieren mirror is limited to its 100 cm diameter, a second series of tests were conducted using the BOS method with musicians from the *Deutsches Nationaltheater und Staatskapelle Weimar*, Germany. This allowed capturing of the airflow to its full extent and to confirm the results which had been measured previously with the schlieren mirror. Additionally, in the second series of tests, air velocity measurements were conducted using omnidirectional hot-wire anemometry with an accuracy of ±1.5% of the measured value, a measurement range of 0.01– 1 *m*/*s*, and a resolution of 0.001 *m*/*s*. The air velocity was recorded over a measurement period of approximately 15 seconds (playing one note) and approximately 30 seconds (playing phrases of a musical piece or improvising) with a sampling interval of 1 second each; the collected values were then averaged. Only three sensors were used to not alter the flow at distances of 20, 52.5, and 85 cm in front of the instrument or mouth. A minimum possible distance of 20 cm was used to avoid possible damage to the sensor from unexpected movement of the subject. From the schlieren mirror images, we concluded that 85 cm is a suitable distance for the farthest sensor. For each instrument, several measurements were conducted (bell, tone holes, mouthpiece, single notes, and musical piece/improvisation).

## 3. Results

In the following sections, the figures displayed in a round shape are recordings of the musician in front of the round schlieren mirror. When the schlieren mirror images showed that the breathing air escapes beyond the 100 cm diameter range of the mirror, an evaluation of the BOS method is presented in rectangular figures. Thus, the following sections show the maximal range of the escaping breathing air for each instrument or singer.

Furthermore, it should be taken into consideration that each musician has developed their blowing technique, which also differs due to their physical characteristics. To control the breathing when playing a wind instrument or singing, several parameters have to be taken into consideration to accomplish stable breath support and, therefore, a clear sound. The breath and its involved organs (throat, larynx and vocal cords, and lungs and diaphragm) form a flexible system. The different wind instruments require specific demands on the blowing techniques. When playing flute instruments, the breathing pressure is even and relatively low. The breathing muscles mostly counteract the passive exhalation forces of the rib cage, which causes the diaphragm to tense during exhalation. When playing brass instruments like the trumpet or the horn, a much higher breathing pressure is needed to overcome the resistance of the mouth lips and the instruments, whereby the abdominal wall muscles are more important. Additionally, the individual physique of the musician plays a major role in the function and process of respiration (Spahn et al. 2013). The angle at which the instrument is held also has to be taken into account to properly estimate the direction in which the air escapes from the mouth, bell, or tone holes as well as when exhaling intermittently between phrases (e.g., oboe, bassoon) or leaking air near the mouthpiece (e.g., clarinet, bass clarinet). Thus, the results presented in this study should not be generalized to all singers and players of wind instruments.

### 3.1. Woodwind instruments

In the case of woodwind instruments, in addition to the breathing air escaping from the bell, the air exiting from the tone holes and the air blown over the mouthpiece in the case of flutes have to be considered. The intermittent exhalation in the case of the oboe and the bassoon, as well as the air leaking near the mouthpiece in the case of the clarinet and the bass clarinet also have to be taken into account.

When looking at the foot joints of the instruments, it can be seen that, especially for small bores, the air escapes laminarly from the bell. After a certain distance, the airflow becomes turbulent and finally mixes with the surrounding room air. How far the air escapes into the room varies concerning the instrument.

#### 3.1.1. Oboe

The tone holes of the oboe are covered with keys that have small holes from which a small amount of air can escape. However, this airflow is hard to capture in the images due to its small range and the small density gradient. Most of the air blown into the instrument escapes from the bell and moves up to 26 cm into the room when playing the note *Bb*3 ≈ 233 *Hz* (Fig. 4a) and up to 30 cm when playing the note *D*6 ≈ 1175 *Hz* (Fig. 4b). Due to the small double reed, in which the air is blown into the instrument, the oboe is played with little breathing air but with high breath pressure. Therefore, the breathing air cannot escape completely from the tone holes and the bell and the oboe player has to exhale intermittently through the mouth and nose between phrases. The air spreads approximately 60 cm into the room (Fig. 4c) but does not exceed the length of the oboe.

**Fig. 4.**
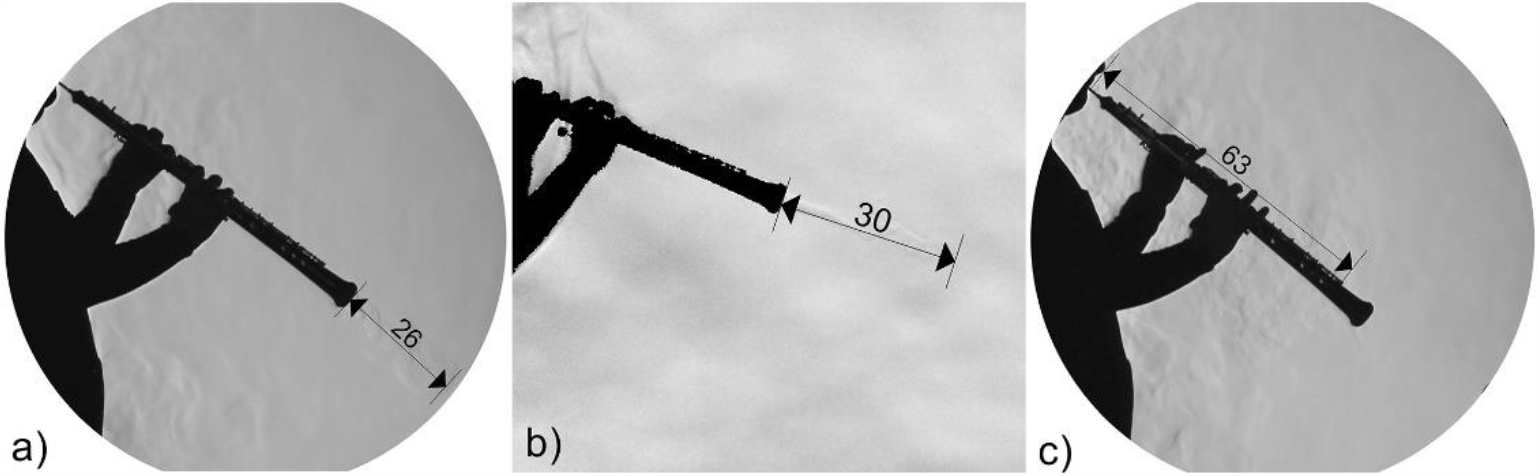
Maximal range of the air escaping from the bell of the oboe while playing the note Bb3 ≈ 233 Hz (a), note D6 ≈ 1175 Hz (b), and when intermittently exhaling (c) [cm]

The anemometry measurements show that the velocity of the breathing air is the highest when playing at high pitches (up to *ν*_*average*_ ≈ 0.09 *m*/*s*, Fig. 5). This confirms the results shown in Fig. 4b, where the air spreads the farthest when playing the note *D*6. High velocities can also be found during intermittent exhalation, followed by a continuous decrease (Fig. 5). Here, the small velocities near the farthest sensor may occur due to the surrounding room air moving since the air spreads only approximately 60 cm when exhaling intermittently (Fig. 4c). When playing at low pitches or playing longer phrases, the velocity of the breathing air escaping from the bell is much lower and, again, decreases with increasing distance.

**Fig. 5.**
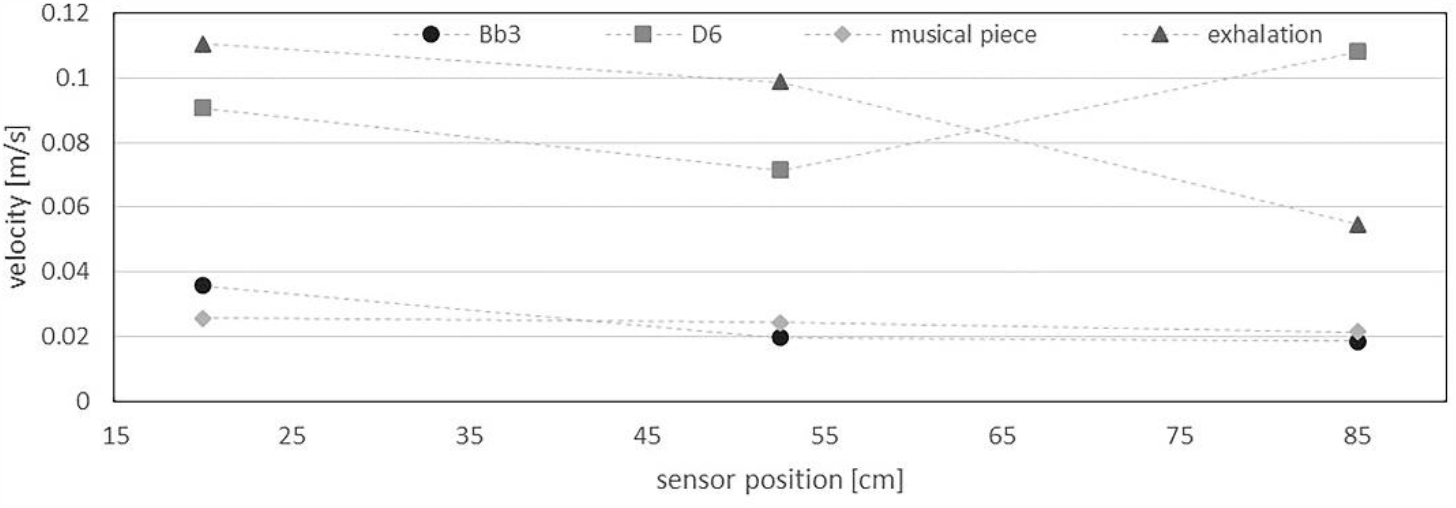
Average velocity of the air escaping from the bell of the oboe (note D6, Bb3, musical piece) and when intermittently exhaling

However, when playing a musical piece, the flow is notably transient (Fig. 6). Air movement of v ≈ 0.02 *m*/*s* at 85 cm from the bell still can be observed. This does not necessarily indicate air movement from the flow escaping the bell; it might be explained by the convective flow of the surrounding room air. The velocity measured by the nearest sensor (at 20 cm from the bell) peaks at *t* ≈ 45 *s*. This corresponds to the large emission of breathing air shown in Fig. 4a and b. These jets are highly transient and escape the bell irregularly.

**Fig. 6.**
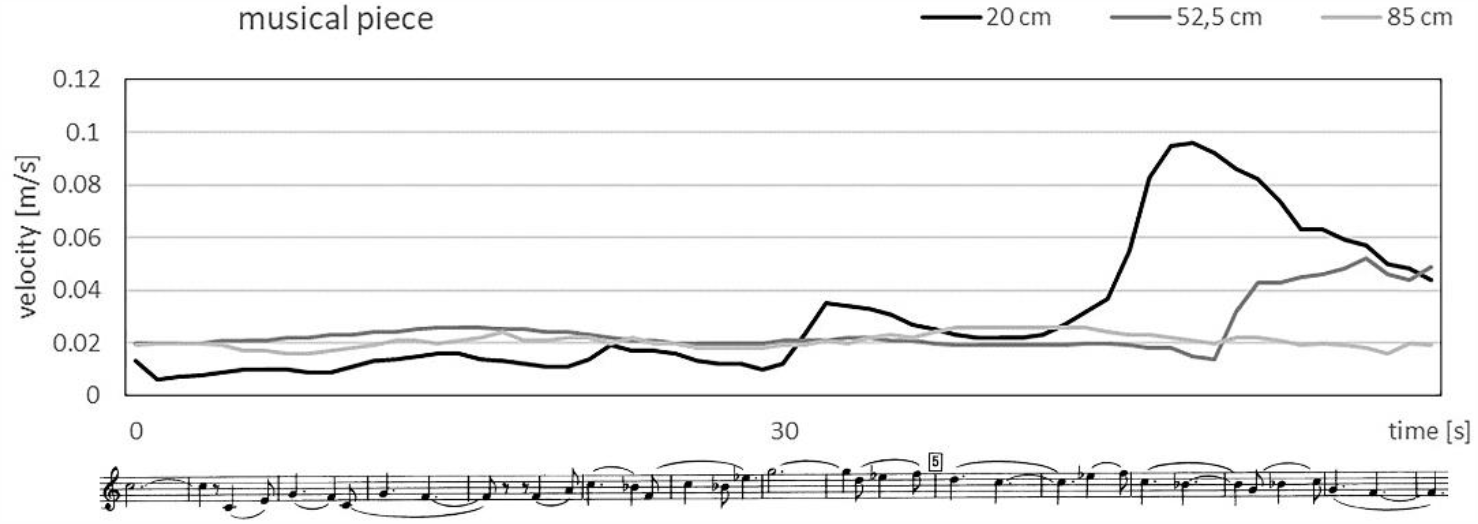
Velocity of the air escaping from the bell of the oboe when playing Bohuslav Martinů: Concerto for Oboe and Small Orchestra

#### 3.1.2. Bassoon

When playing the bassoon at lower pitches where most of the tone holes are covered, nearly all the air blown into the instrument escapes from the bell (Fig. 7a). At high pitches, where many tone holes are open, hardly any air escapes from the bell. Here, a smaller jet can be observed near the tone holes. The largest airflow escaping from the tone holes occurs when playing the note *F*3 ≈ 175 *Hz*, where most of the tone holes are not covered (Fig. 7b). Similar to the oboe, the bassoon is played at high breath pressure. Due to the small double reed, from which the air is blown into the instrument, the air cannot escape completely from the bell and the tone holes. Thus, it needs to be exhaled intermittently and, therefore, spreads approximately 50 cm into the room (Fig. 7c).

**Fig. 7.**
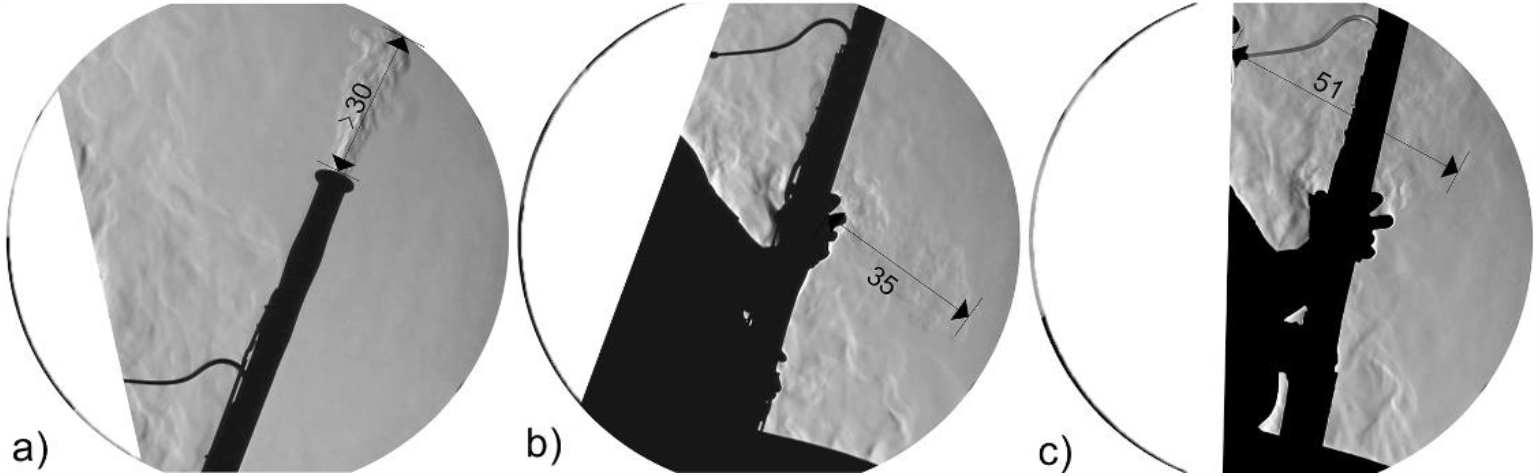
Maximal range of the air escaping from the bell or the tone holes of the bassoon while playing the note Bb1 ≈ 58 Hz (a), note F3 ≈ 175 Hz (b), and when intermittently exhaling (c) [cm]

Investigations with the BOS method showed that the airflow escaping from the bell does not move any farther than what the schlieren images illustrated (Fig. 7a) and, thus, confirm the measurements with the schlieren method.

The anemometry measurements show that the highest velocity of the air escaping from the bell up to approximately 0.13 *m*/*s* occurs when playing the note Bb1, where nearly all the air escapes from the bell (Fig. 8). When playing at high pitches, nearly no velocity of the air escaping from the bell could be measured. The velocity of the air escaping from the tone holes when playing the note F3 increases slightly but continuously with increasing distance. Once again, intermittent exhalation features the highest air velocities similar to the oboe.

**Fig. 8.**
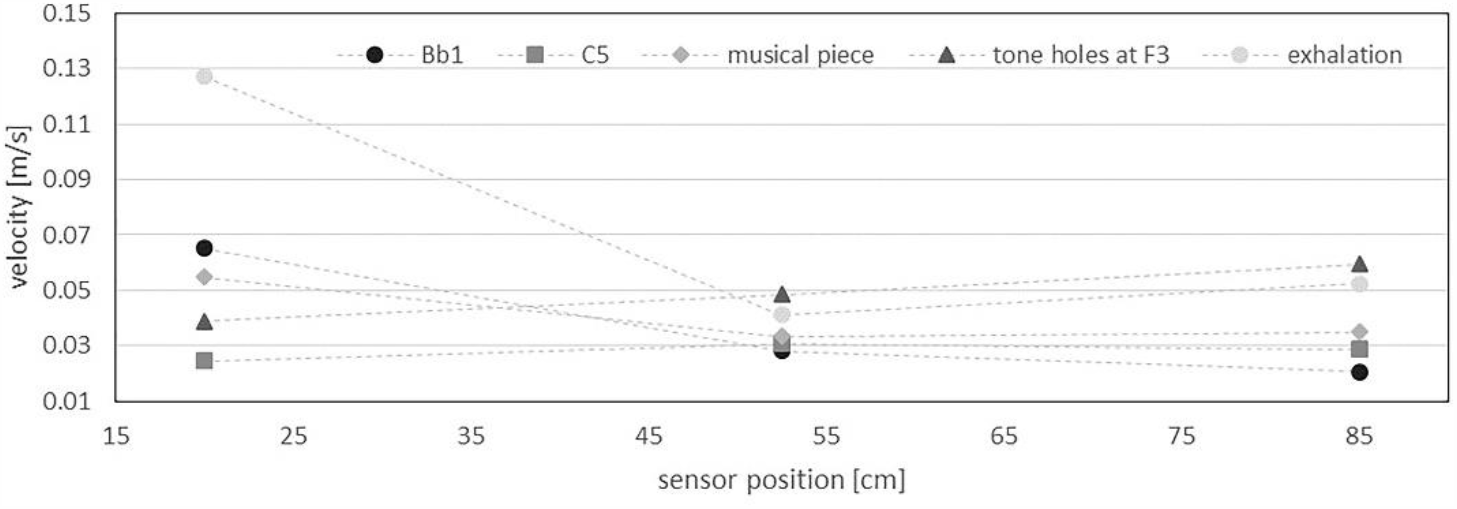
Average velocity of the air escaping from the bell of the bassoon (single note Bb1, C5, musical piece), from the tone holes (F3), and when intermittently exhaling

#### 3.1.3. Bb clarinet and bass clarinet

The breathing air, which is blown into the instrument, can escape completely from the bell and the tone holes and reaches up to approximately 25 cm from the bell into the room when playing at low pitches (Fig. 9a) and slightly more when playing at high pitches (Fig. 9b). At the tone holes, which are covered with keys, almost no moving air can be observed due to its small range and small density gradients. In addition to the air escaping from the bell, the breathing air leaking at the mouthpiece has to be considered. This occurs when the lips of the clarinet player tire during long rehearsals or concerts.

**Fig. 9.**
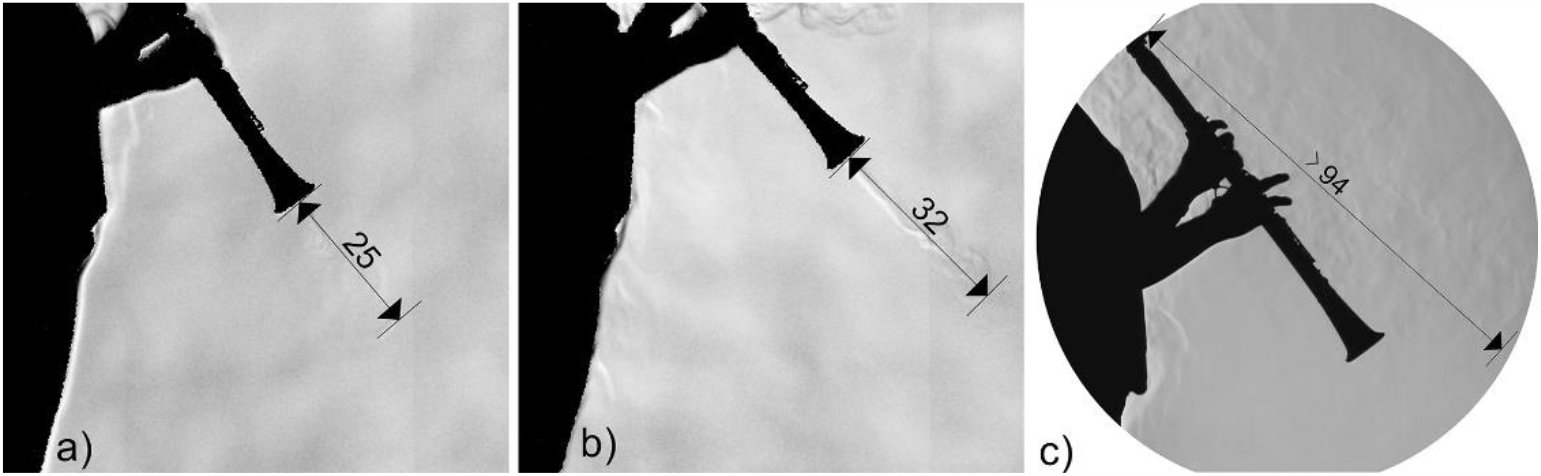
Maximal range of the air escaping from the bell of the Bb clarinet while playing the note C#3 ≈ 139 Hz (a), note E6 ≈ 1319 Hz (b), and the air leaking at the mouthpiece (c) [cm]

Leaking air can also be observed when playing a tone with an accent (staccato) or with untrained musicians playing the clarinet, such as beginners or older clarinet players. Here, the clarinet player simulated her lips tiring, which results in the air leaking near the mouthpiece (Fig. 9c).

Investigations with the BOS method showed that the airflow leaking at the mouthpiece does not move any farther than what the schlieren images illustrated and, therefore, confirm the measurements with the schlieren method.

The anemometry measurements show that the highest velocity of the air escaping from the bell occurs when playing at low pitches opposite to the visualization in Fig. 9a and b. The air leaking at the mouthpiece has a slightly higher velocity up to approximately 0.15 *m*/*s*, which decreases rapidly until it is almost immeasurable at the farthest sensor (Fig. 10).

**Fig. 10.**
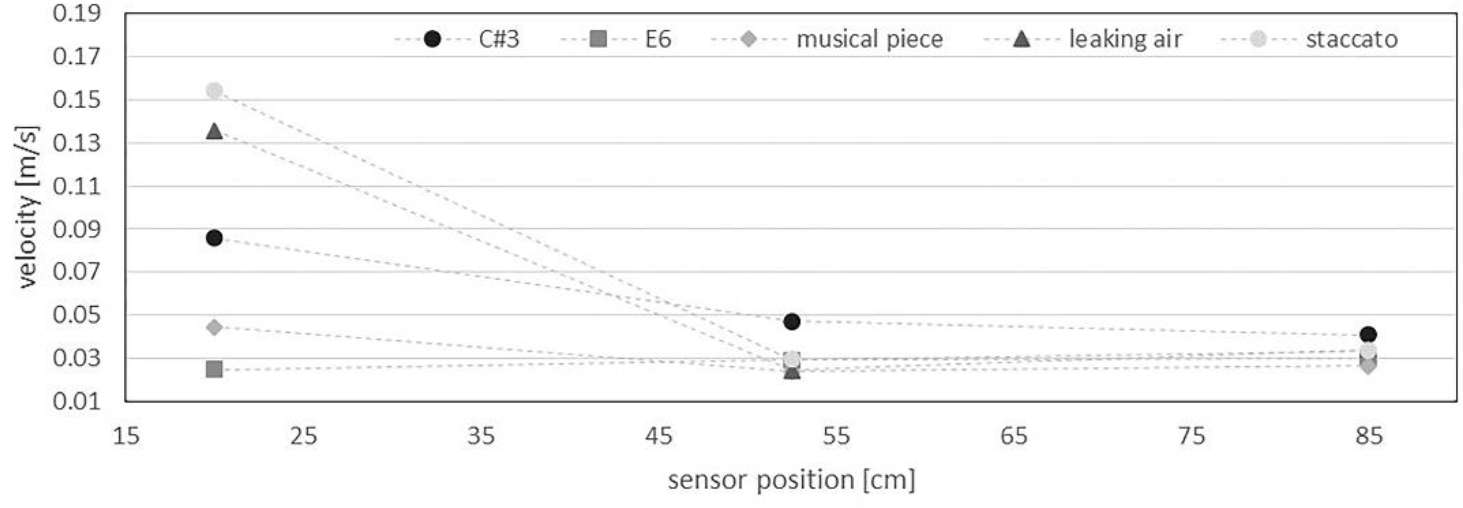
Average velocity of the air escaping from the bell of the Bb clarinet (single note C#3, E6, musical piece) and near the mouthpiece (leaking air, staccato)

The air escaping from the bell of the bass clarinet is transported nearly vertically into the room when the instrument was aligned vertically. Similar to the Bb clarinet, almost no jet of air can be seen at the tone holes. When playing at lower pitches where most of the tone holes are covered, most of the air escapes from the bell of the instrument (Fig. 11a). When playing a musical piece, the air reaches the farthest into the room with a distance of up to 33 cm (Fig. 11c).

**Fig. 11.**
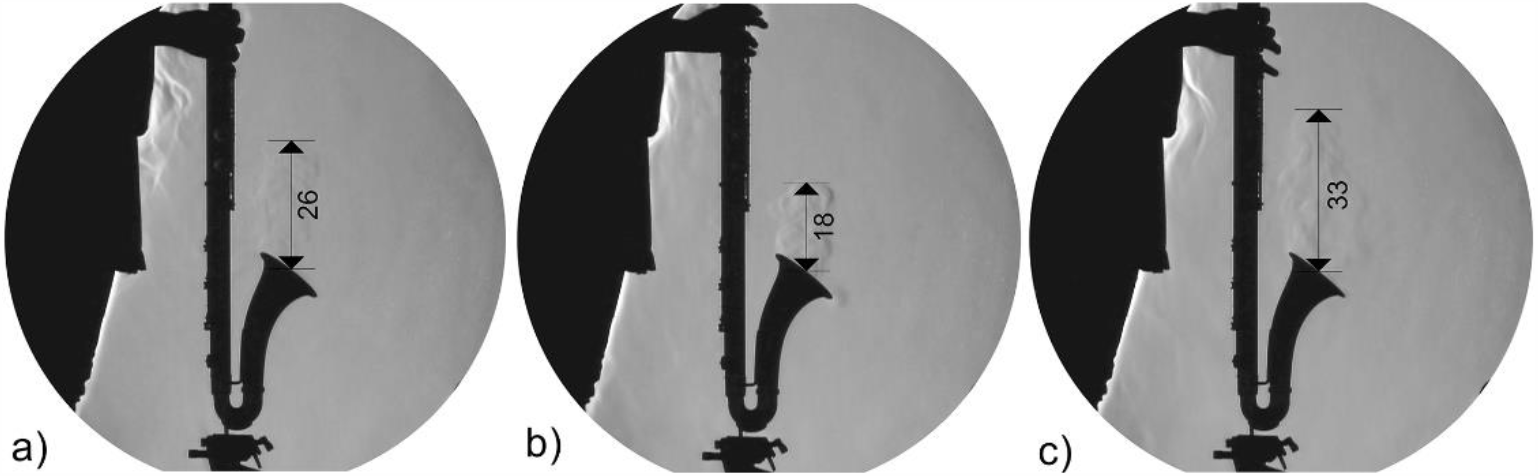
Bass clarinet while playing the note D2 ≈ 73 Hz (a), note Bb4 ≈ 466 Hz (b), and the maximal range of the air escaping from the bell (c) [cm]

#### 3.1.4. Grand flute, piccolo, and alto flute

The air escaping from the foot joint of the grand flute and the piccolo is similar. When playing these instruments, a similarly large jet of air escapes from the end of the flute with a range of approximately 20 cm (Fig. 12a and b). Regarding the alto flute, the air escapes much farther into the room compared to the smaller flutes (Fig. 12c). Near the tone holes of the instruments, which are covered with keys, hardly any escaping air can be observed.

**Fig. 12.**
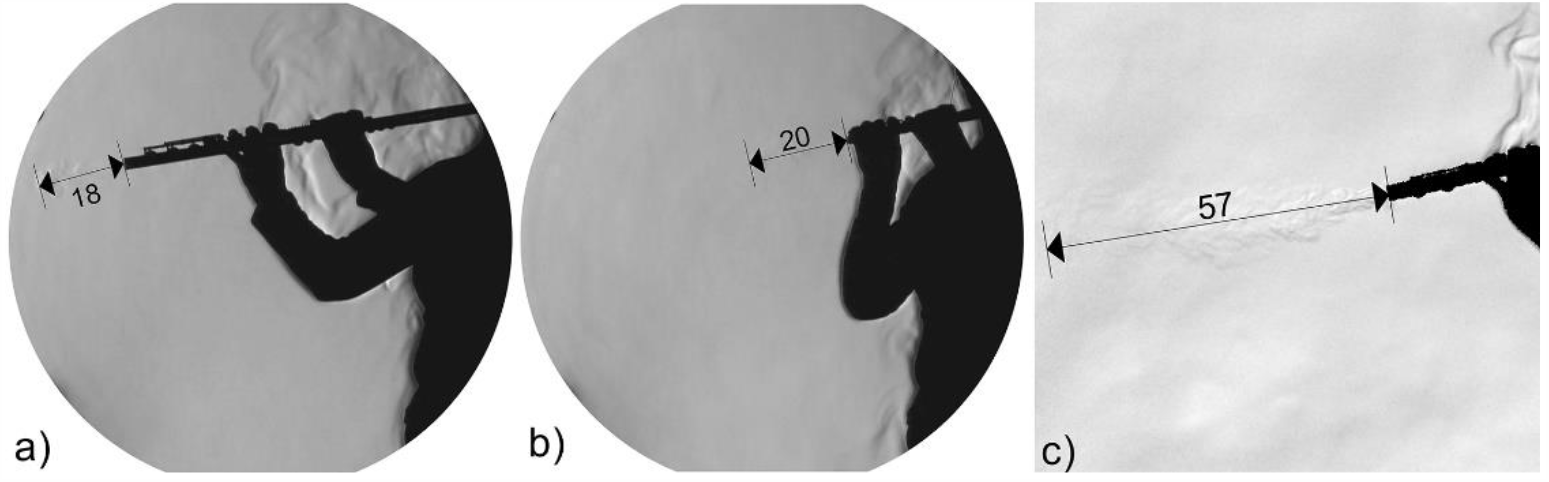
Maximal range of the air escaping from the foot joint of the grand flute while playing the note E4 ≈ 330 Hz (a), of the piccolo while playing the note E5 ≈ 659 Hz (b), and of the alto flute when playing the note G2 ≈ 98 Hz (c) [cm]

Within the range of each instrument, the air being expelled through the foot joint escapes the farthest when playing at low pitches, where most of the tone holes are covered (Fig. 12 and Fig. 13).

**Fig. 13.**
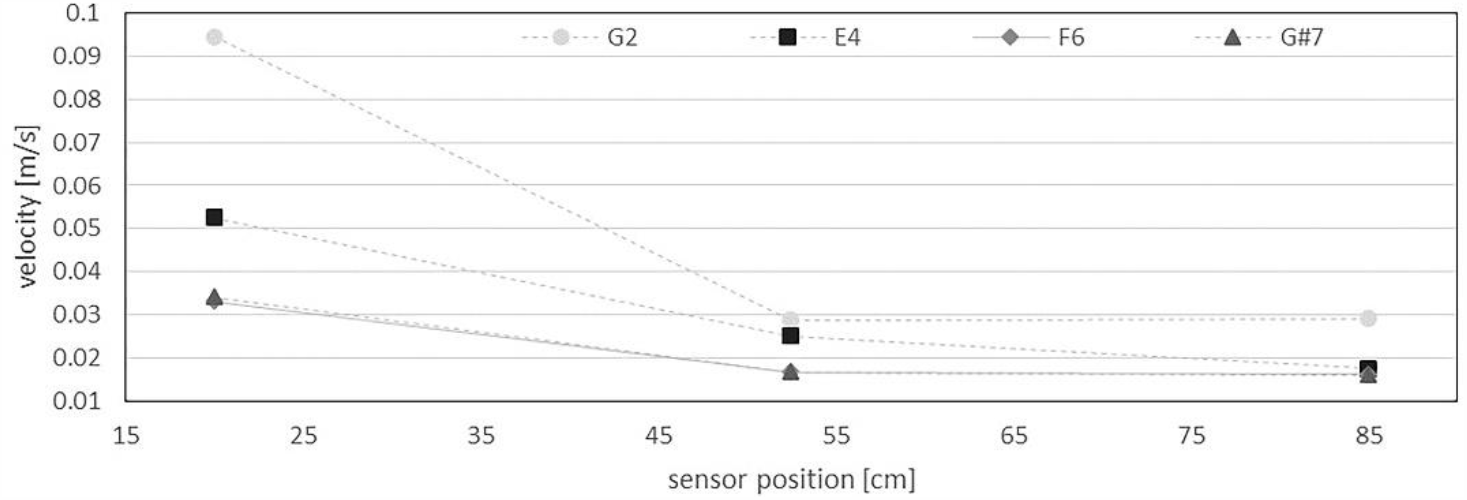
Average velocity of the air escaping from the foot joint (single note G2 (alto), E4 and F6 (grand flute), and G#7 (piccolo))

Fig. 13 shows that the velocity of the air escaping from the foot joint when playing the alto flute at low pitches is the highest (up to approximately 0.1 *m*/*s*) compared to the smaller flutes playing at higher pitches (approximately 0.05 *m*/*s* for the grand flute and 0.03 *m*/*s* for the piccolo). This also confirms the BOS image of the alto flute in Fig. 12c, where the air escapes the farthest from the foot joint of the alto flute.

However, most of the breathing air is blown over the mouthpiece of the flutes. The air is cut at the splitting edge and moves either into the bore of the instrument or into the room over the mouthpiece. Here, the air escapes the farthest into the room (Fig. 14) compared to the air escaping from the foot joints (Fig. 12).

**Fig. 14.**
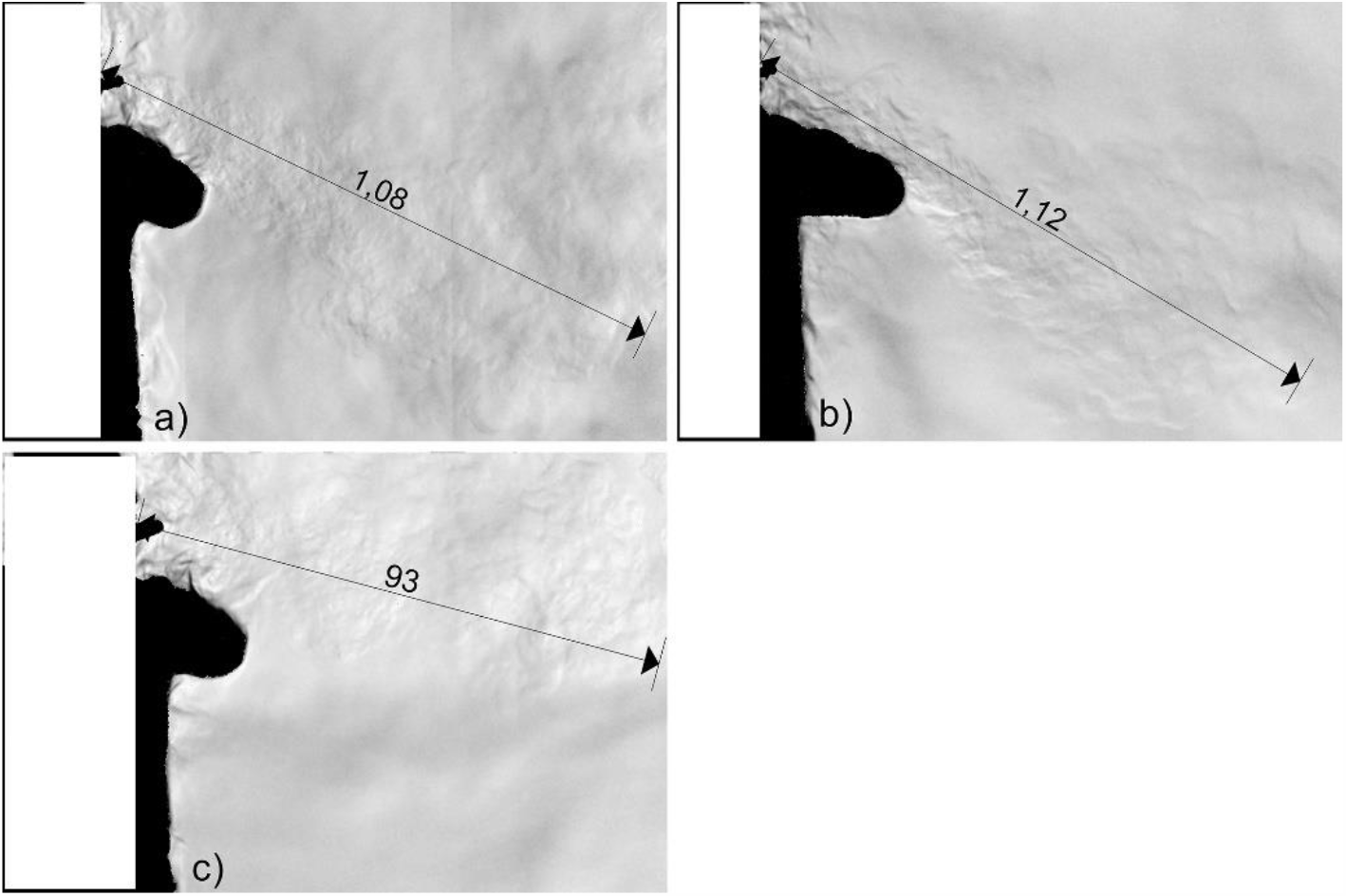
Maximal range of the air blown over the mouthpiece of the grand flute (a), the piccolo (b), and the alto flute (c) when playing a musical piece at medium pitches [cm]

When playing at low pitches, the flute player blows more air directly into the mouthpiece of the instrument compared to high pitches, where most of the air is not blown into the instrument but over its mouthpiece. Furthermore, when playing notes in forte, more air is blown at a higher velocity over the mouthpiece of the instrument. To keep the pitch stable, the angle at which the air is blown into the instrument is varied (Spahn et al. 2013). The evaluations show that the higher the instrument and therefore the tone played, the more air is blown over the mouthpiece in a vertical direction (Fig. 14b). Regarding the alto flute, which can be played at the lowest pitches in relation to the grand and piccolo, the air is blown nearly horizontally into the room (Fig. 14c). Additionally, the air blown over the mouthpiece moves farther when playing the piccolo at high pitches (up to approximately 0.11 *m*/*s*) compared to playing the grand and alto flute at lower pitches (Fig. 15). Furthermore, the velocity of the air blown over the mouthpiece increases when playing at high pitches, which confirms the visualizations in Fig. 14.

**Fig. 15.**
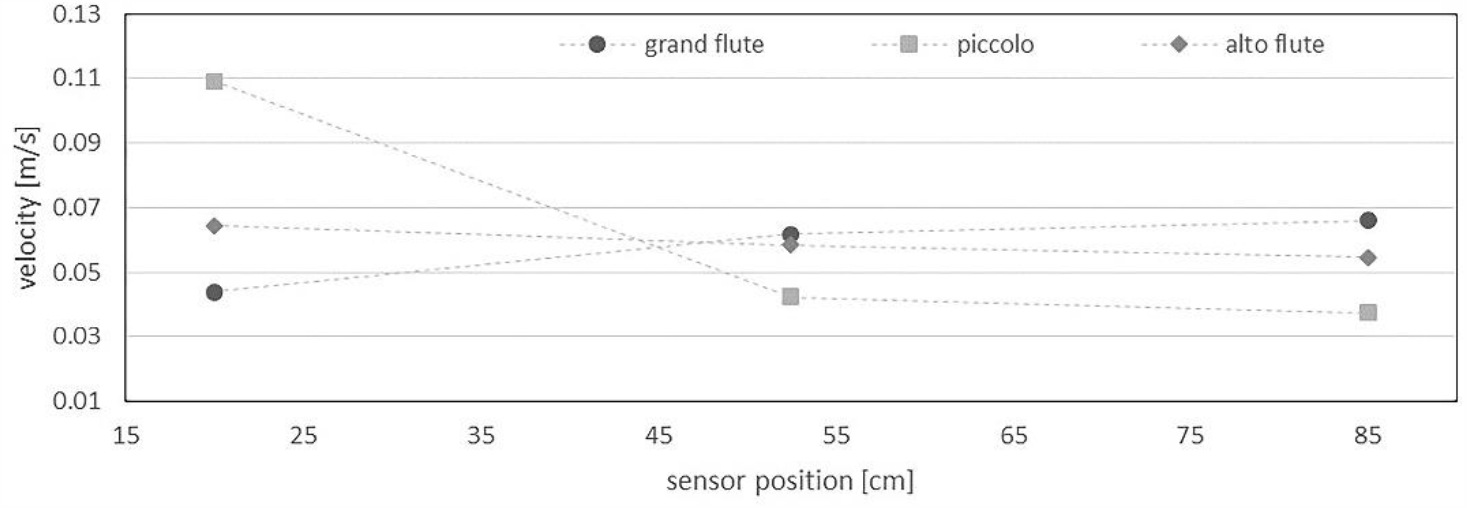
Average velocity of the air blown over the embouchure when playing a musical piece at medium pitches Brass instruments

### 3.2. Brass instruments

The breathing air blown into the mouthpiece of the brass instruments can escape completely from the bell. Usually, no air leaks near the mouthpiece. Leaking air can only be observed with beginners, who are not assessed in this study. The various natural tones of a brass instrument are played by varying the pressure of the lips and the breathing air. This affects the range and angles at which the air exits the bell. If non-natural tones are played, valves (e.g., Bb trumpet, French horn, F tuba) or slides (e.g., tenor trombone) are used to lengthen or shorten the air column inside the instrument to be able to play at every possible pitches and scales.

In contrast to the woodwind instruments, the air escaping from the bells of the brass instruments is highly turbulent. This is due to the larger diameter of the bells compared to the foot joints of the woodwind instruments. Moreover, mutes and dampers are used commonly with brass instruments. When playing with mutes or dampers, the air escapes through the narrow gap between the brass of the instrument and the material of the damper. This alters the spread of the air escaping from the bell significantly. Further details about the impact of dampers are presented and discussed for each instrument in the following sections.

#### 3.2.1. Bb trumpet

The pitch of the played note affects how far and at which angle the air escapes from the bell (Fig. 16a and b). When playing at high pitches, the air escapes at a higher velocity and higher pressure from the mouth of the trumpet player. However, when playing at high pitches and high volume, no larger volume of air is required (Spahn et al. 2013). If dampers are used, the airflow escaping from the bell is restricted significantly (Fig. 16c).

**Fig. 16.**
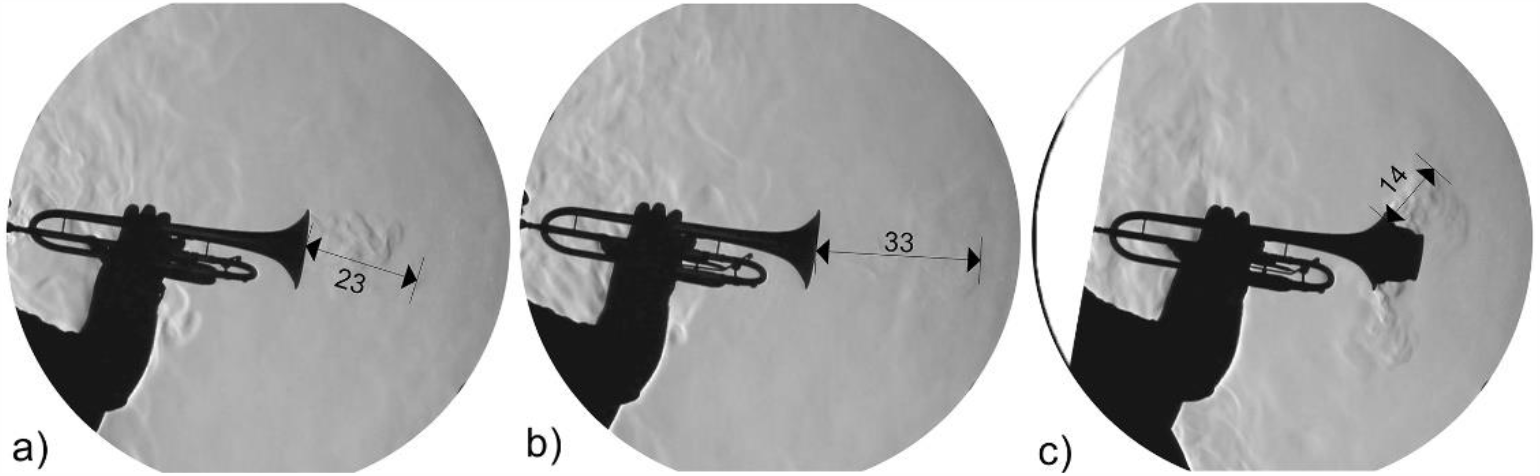
Maximal range of the air escaping from the bell of the Bb trumpet while playing the note Bb3 ≈ 233 Hz (a), note Bb5 ≈ 932 Hz (b) and with cup mute (fully closed) (c) [cm]

When playing at lower pitches or playing a musical piece, only a low velocity of the escaping airflow can be measured at a distance of approximately 20 cm in front of the bell (Fig. 17). When playing at high pitches, the air escapes at a higher velocity up to approximately 0.07 *m*/*s*, which corresponds to the visualizations in Fig. 16b. When examining the values measured by the sensor that is the farthest from the bell at 85 cm, the measurements show little air movement mainly resulting from the movement of the surrounding indoor air.

**Fig. 17.**
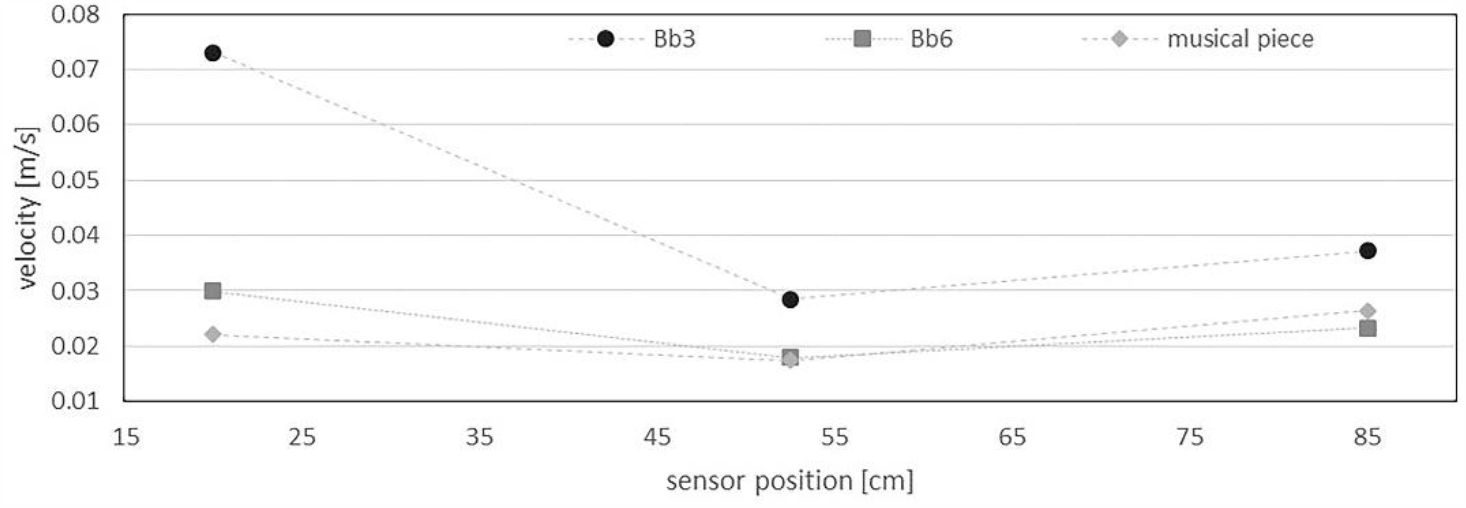
Average velocity of the air escaping from the bell of the Bb trumpet (single note Bb3, Bb6, musical piece)

#### 3.2.2. Tenor trombone

The pitch of the note played affects how far and at which angle the air escapes from the bell (Fig. 18a and b). If dampers are used, the airflow escaping from the bell is restricted significantly (Fig. 18c). Unlike other brass instruments, the notes that vary from the natural tone scale of the instrument are produced by moving the slide of the trombone, thereby lengthening the air column inside the bore of the instrument. Hence, the air escaping from the bell is swirled when playing non-natural tones.

**Fig. 18.**
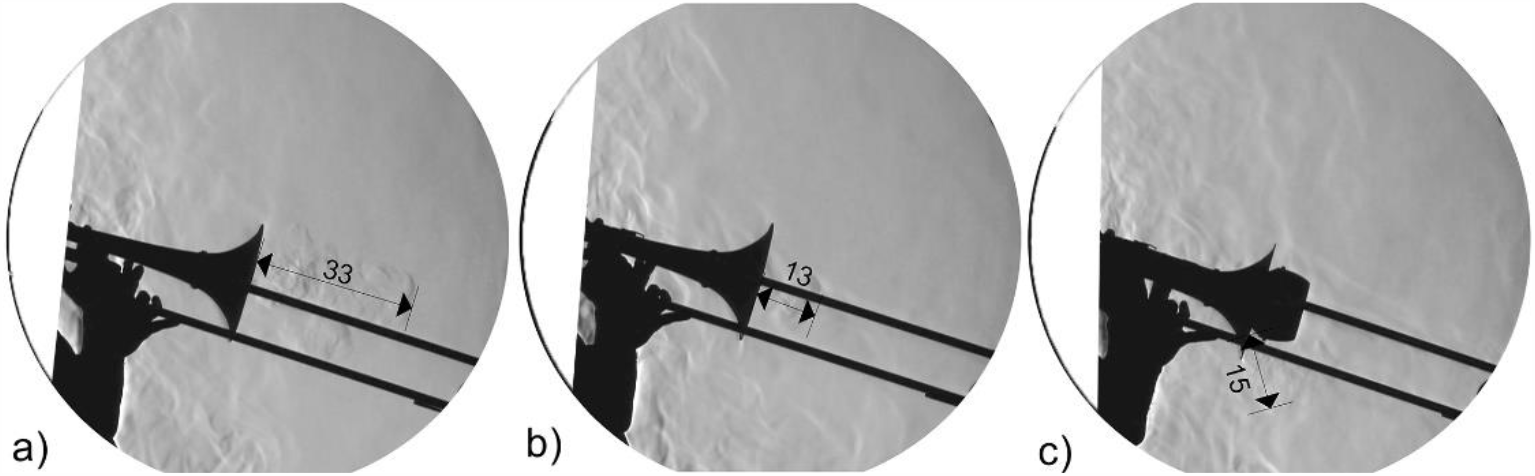
Maximal range of the air escaping from the bell of the tenor trombone while playing the note Bb1 ≈ 58 Hz (a), note Bb3 ≈ 233 Hz (b), and with damper (c) [cm]

Fig. 19 shows that the velocity of the escaping breathing air is generally low with the highest value when playing at high pitches (up to approximately 0.05 *m*/*s*). For playing at lower pitches and a musical piece, the velocity decreases with increasing distance (the final increase is the result of surrounding convective flows). When playing the note Bb1, the velocity of the escaping breathing air decreases constantly. However, compared to the other brass instruments, the velocity of the breathing air escaping from the bell is rather low.

**Fig. 19.**
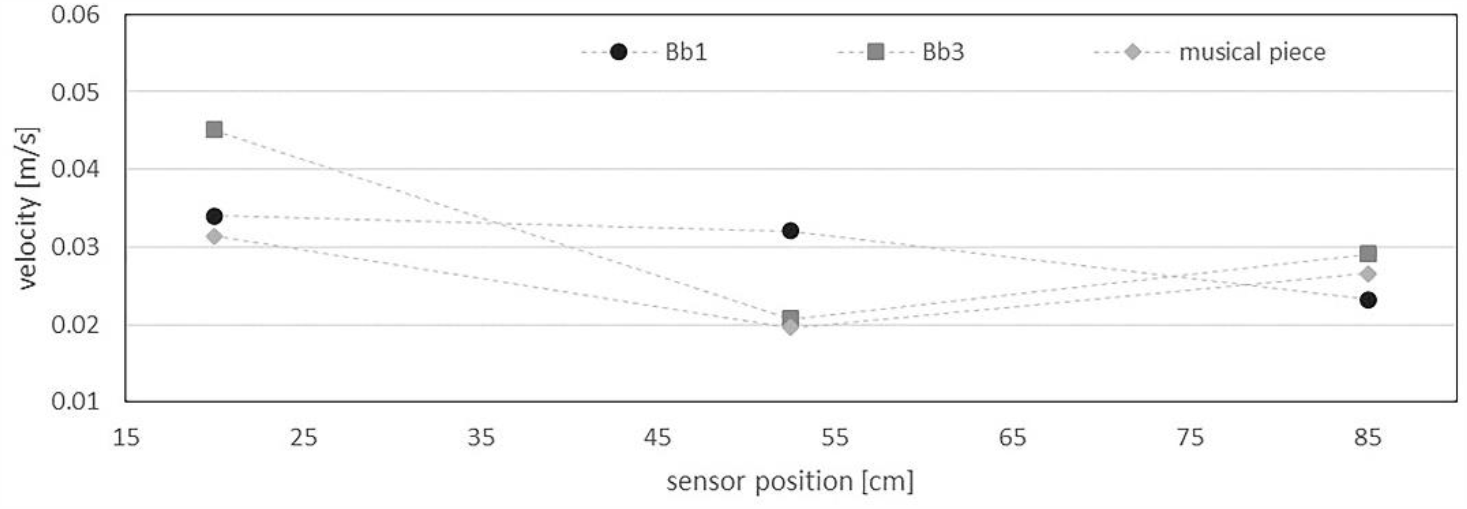
Average velocity of the air escaping from the bell of the tenor trombone (single note Bb1, Bb3, musical piece)

#### 3.2.3. French horn

The French horn is usually held with the right hand; the valves are controlled with the left hand. When playing at higher pitches, the air escapes farther into the room compared to playing at lower pitches. The escaping breathing air reaches the farthest when playing a musical piece (Fig. 20a). If dampers are used, the jet of air escaping from the bell is reduced and bent along the right hand or forearm (Fig. 20b). An exception is the stopping mute, where the air escapes from a narrow tube at a higher pressure. Thus, it spreads farther than 70 cm into the room and escapes over the edge of the schlieren mirror. However, the flow is more difficult to detect due to the lower density gradient (Fig. 20c).

**Fig. 20.**
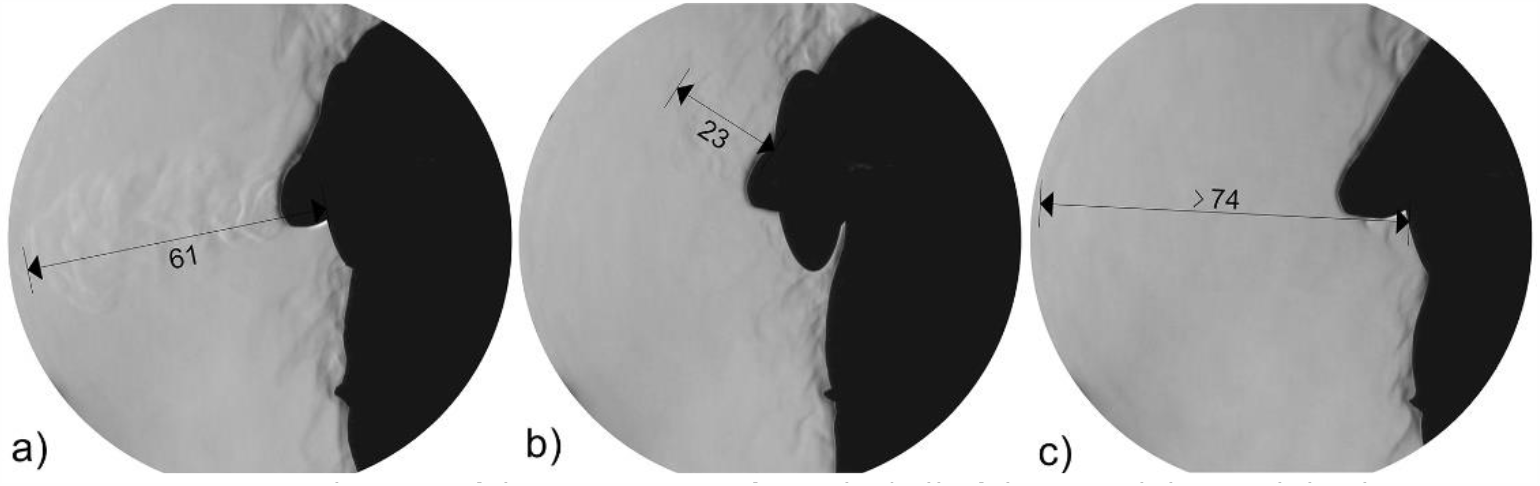
Maximal range of the air escaping from the bell of the French horn while playing a musical piece (a), with damper (b), and stopping mute (c)[cm]

Investigations with the BOS method showed that the air escaping from the bell with stopping mute does not move any farther than what the schlieren images illustrated and, therefore, confirm the measurements with the schlieren method.

Fig. 21 shows that the velocity of the escaping breathing air is ν < 0.03 *m*/*s*. As the measured values are similar to the background air velocity during the measurements without any instruments, they are negligible.

**Fig. 21.**
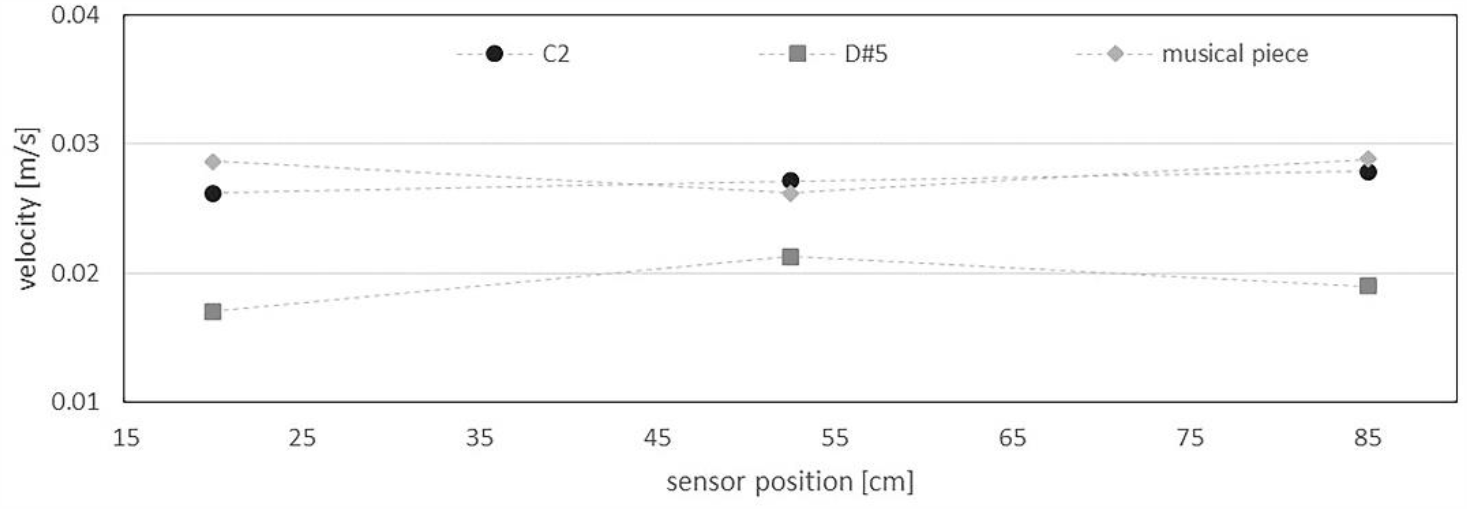
Average velocity of the air escaping from the bell of the French horn (single note Bb1, Bb3, musical piece)

#### 3.2.4. F tuba

Fig. 22a shows that the air escaping from the bell is transported into the room only over a short distance. In contrast to the other brass instruments, the air escapes farther when dampers are used (Fig. 22c).

**Fig. 22.**
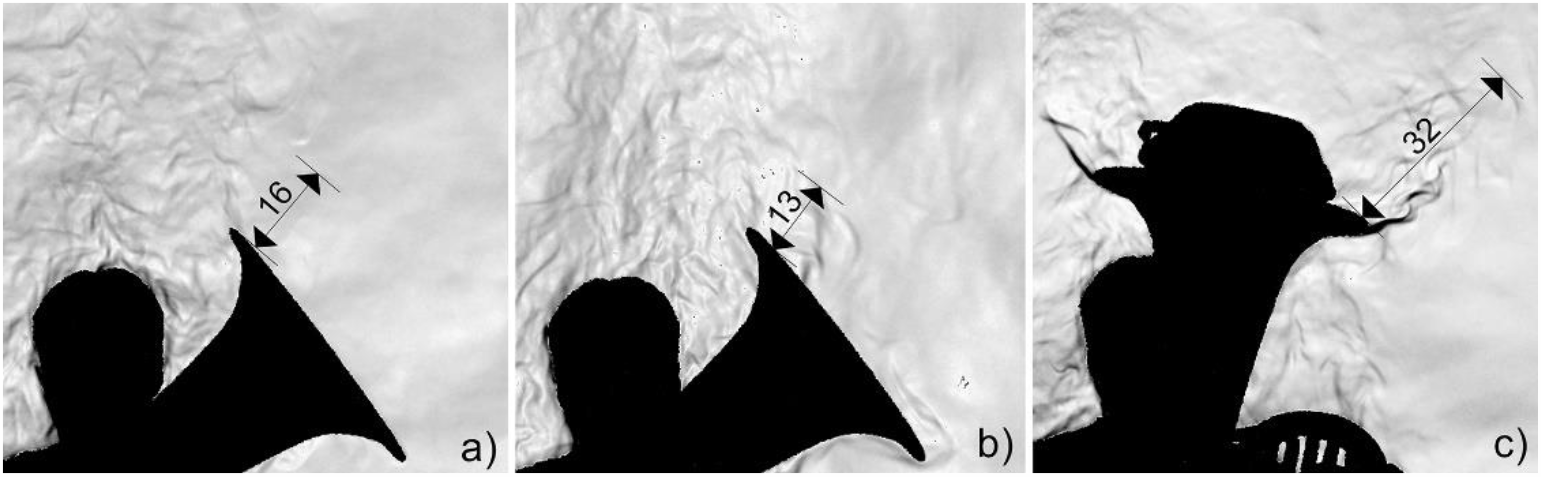
Maximal range of the air escaping from the bell of the F tuba while playing the note F1 ≈ 44 Hz (a), note F4 ≈ 349 Hz (b), and with damper (c)[cm]

The anemometry measurements show that the breathing air escapes at higher velocity when playing at high pitches (Fig. 23). As the airflow only reaches approximately 15 cm into the room, the velocity measured with the farthest sensor only shows random disturbances, which could also occur due to the breathing air escaping from the bell building small eddies. When playing at lower pitches, the air escapes at a low velocity and reaches a constant velocity near the middle sensor at 52.5 cm.

**Fig. 23.**
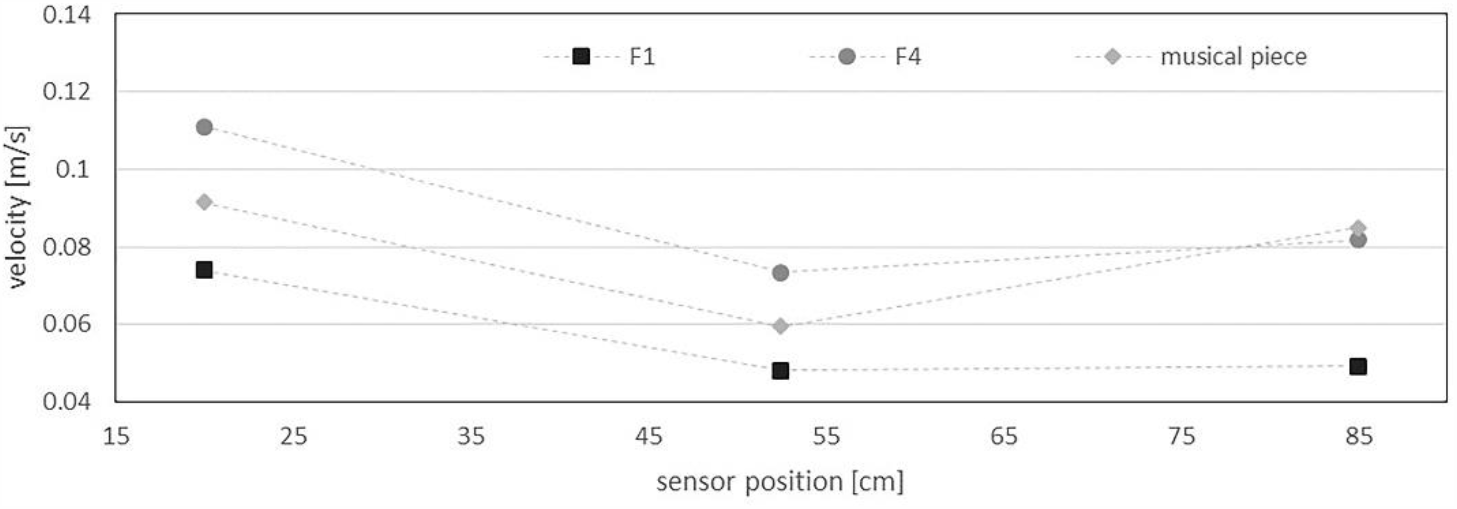
Average velocity of the air escaping from the bell of the F tuba (single note F4, F1, musical piece)

### 3.3. Singers

When singing a note, the greatest air spread can be observed at the beginning of the tone production. If a tone is held or a longer phrase is sung, only a small amount of air escapes from the mouth of the singer. In contrast, the greatest air spread can be seen when singing consonants and with high activities during articulation.

#### 3.3.1. Baritone

Evaluations show that the range of the exhaled air hardly differs in the variation of the pitch (e.g., when singing scales) (Fig. 24a and b). An increasing air spread can be observed when singing demanding musical pieces that include consonants and which require high levels of articulation as well as a strong command of the voice (Fig. 24c).

**Fig. 24.**
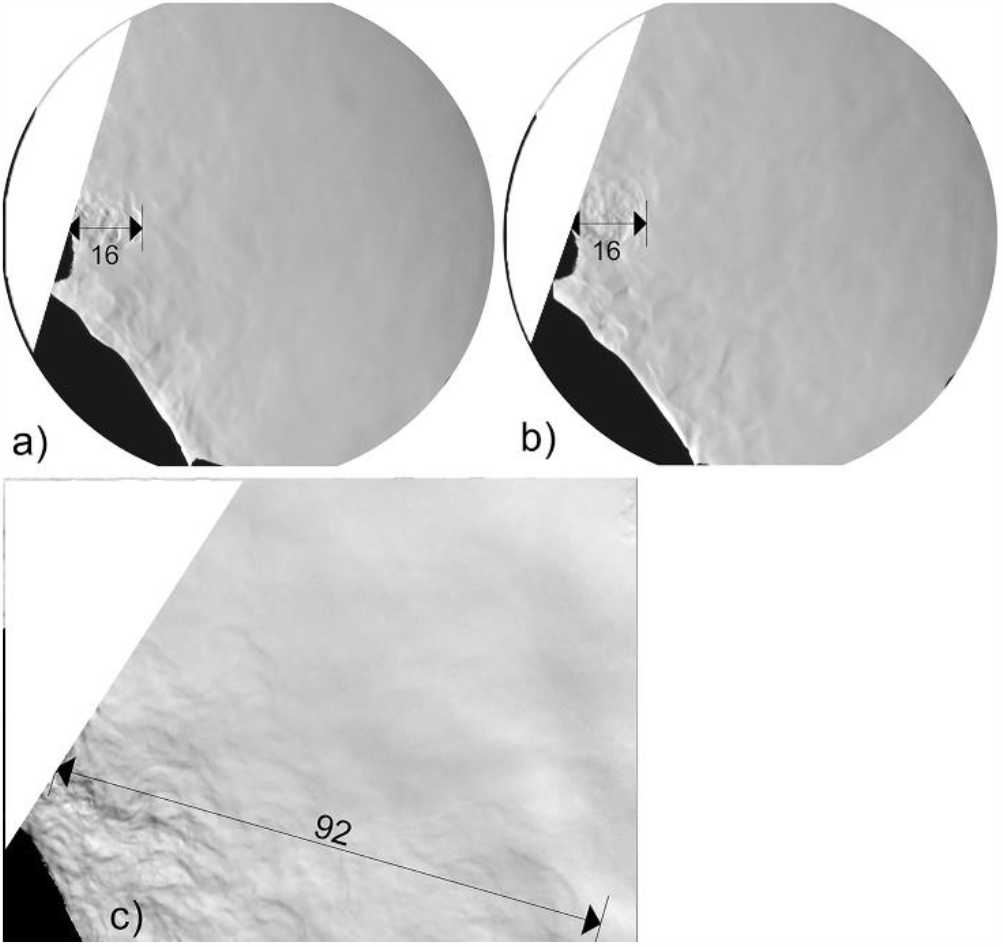
Maximal range of the air escaping from the mouth while singing the note G2 ≈ 98 Hz (a), note D#5 ≈ 622 Hz (b), and a demanding musical piece (c) [cm]

The anemometry measurements show that the velocity of the escaping breathing air is the highest when singing at low pitches or a demanding musical piece with a velocity up to approximately ν < 0.13 *m*/*s*. The air velocity measured when singing a musical piece was still detectable by the farthest sensor at 85 cm from the mouth. This can also be seen in Fig. 24 where the air spreads farther than 90 cm into the room. When singing single notes at either low or high pitches, the velocity cannot be detected near the middle sensor, which corresponds to the visualizations shown in Fig. 24a and b.

#### 3.3.2. Soprano

Similar to the baritone, the evaluations show that the range of the exhaled air hardly differs in the variation of the pitch (e.g., when singing scales) (Fig. 26a and b). Again, an increasing air spread can be observed when singing demanding musical pieces that include consonants and which require high levels of articulation as well as a strong command of the voice (Fig. 26c).

**Fig. 25.**
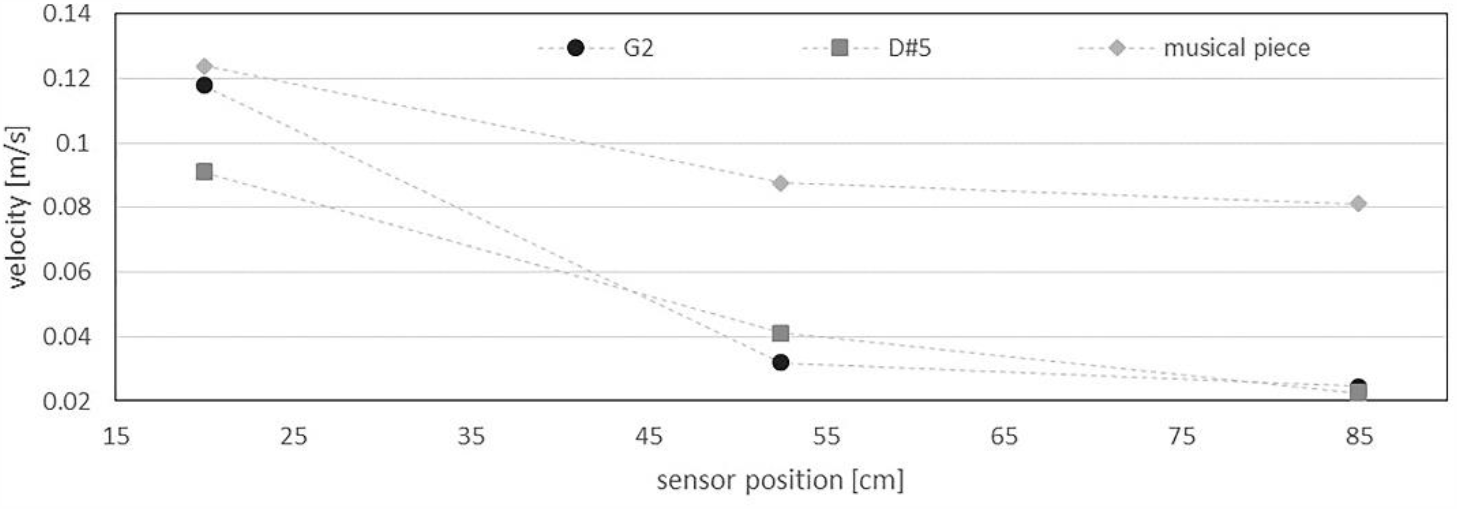
Average velocity of the air escaping from the mouth of the baritone (single note G2, D#5, musical piece)

**Fig. 26.**
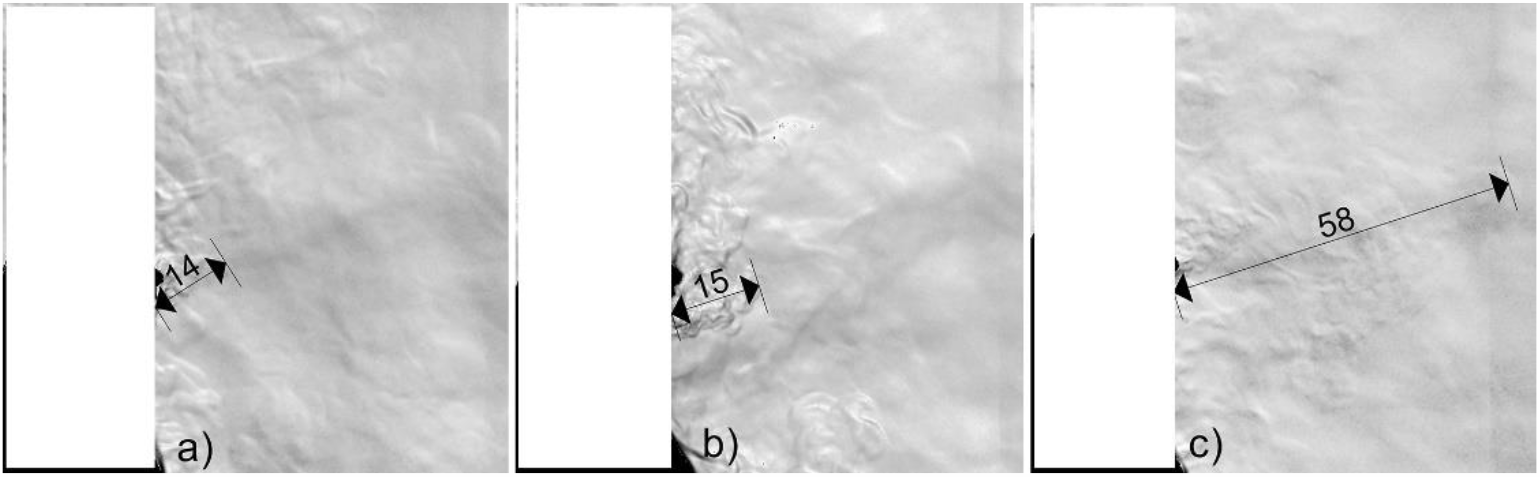
Maximal range of the air escaping from the mouth while singing the note A3 ≈ 220 Hz (a), note A5 ≈ 880 Hz (b), and a demanding musical piece (c) [cm]

Similar to the velocity measurements of the baritone, the velocity of the escaping breathing air from the soprano is the highest when singing a demanding musical piece (up to approximately 0.14 *m*/*s*). However, in contrast to the baritone singer, a lower air velocity can be observed for the soprano when singing at low pitches. Near the middle sensor hardly any air movement can be observed, which corresponds to both singing a single note or a musical piece (Fig. 27).

**Fig. 27.**
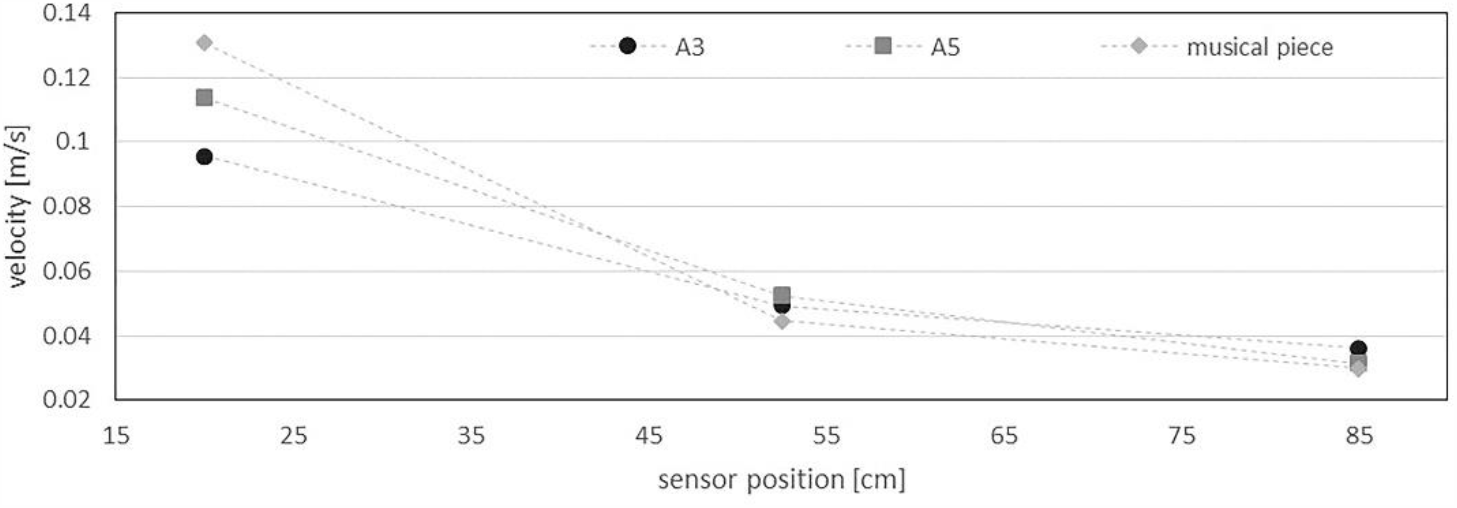
Average velocity of the air escaping from the mouth of the soprano (single note A3, A5, musical piece)

Fig. 28 shows the transient behavior of the air velocity recorded during the singing of an aria. It illustrates that only the nearest sensor (at 20 cm from the mouth) measures values of up to ν ≈ 0.2 *m*/*s*. The sensors that are farther away from the singer only show small velocity values with maximal 0.08 *m*/*s*. However, the velocity measured at the middle sensor (at 52.5 cm) is shifted slightly in terms of chronology compared to the first sensor at 20 cm. This applies to the airflow measured by the first sensors and captured by the second sensor a few seconds later when moving at these low velocities.

**Fig. 28.**
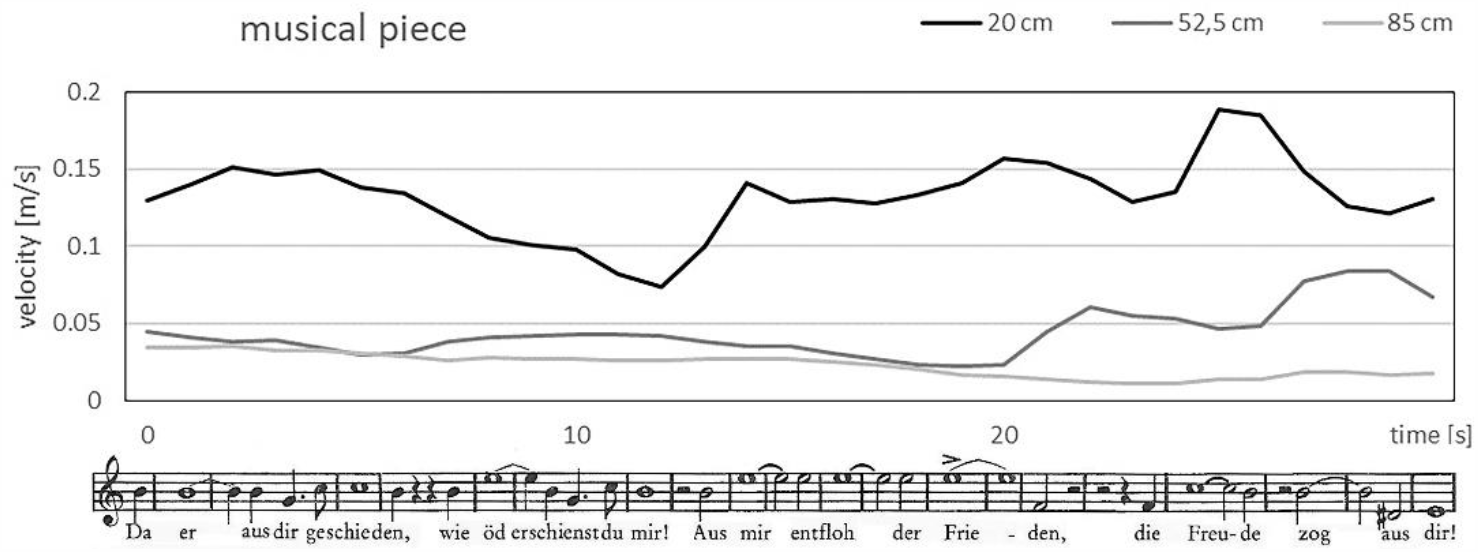
Velocity of the air escaping from the mouth of the soprano singer when singing Richard Wagner: Dich, teure Halle, grüß’ ich wieder. Tannhäuser und der Sängerkrieg auf der Wartburg

## 4. Discussion and outlook

Fig. 29 shows a comparison of the maximal spreading ranges of the escaping breathing air from different instruments and singers. However, not only the spreading range varies strongly but also the angle at which the air escapes into the room.

**Fig. 29.**
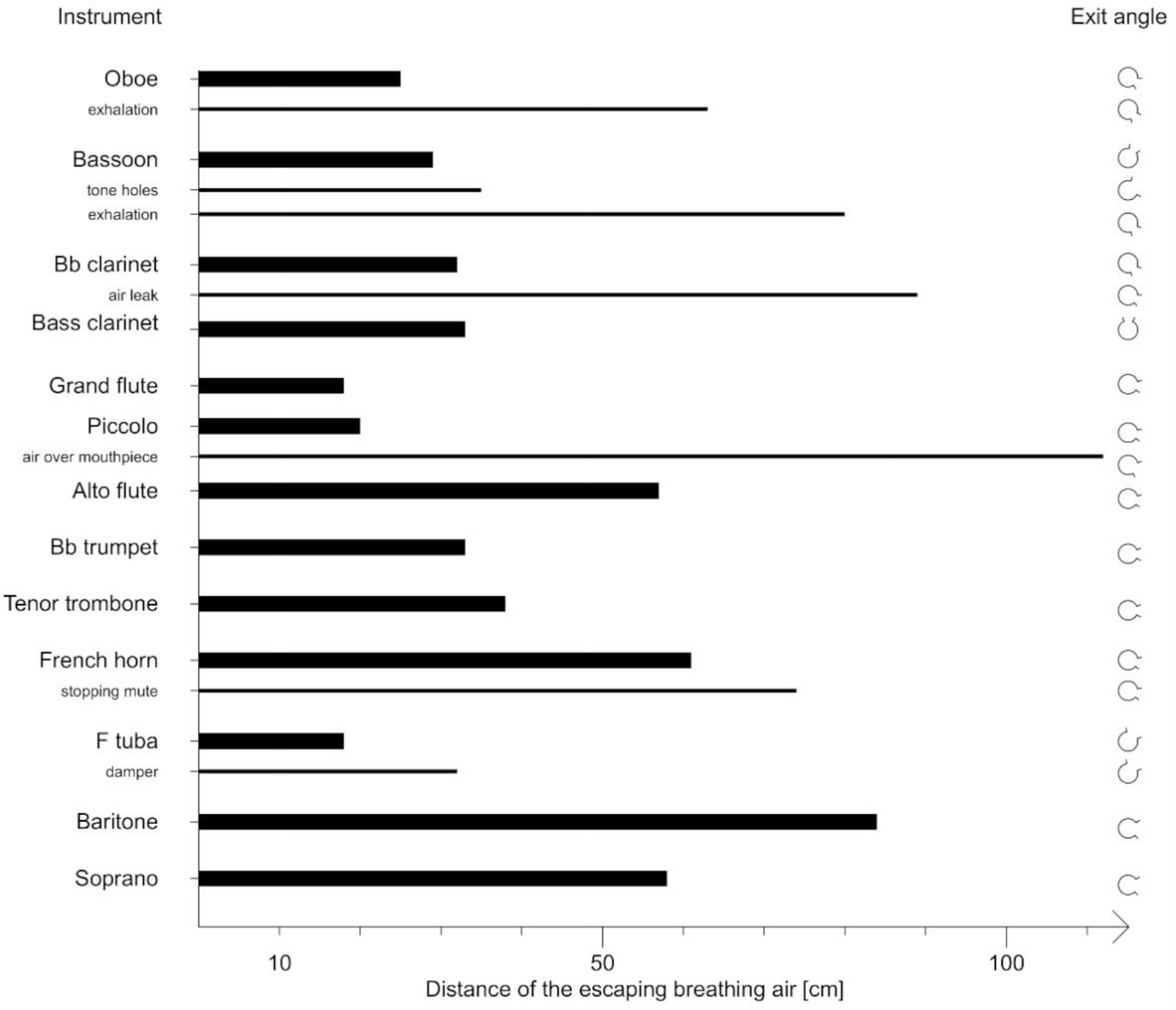
Maximal spreading distance of the breathing air from wind instruments and singers

For woodwind instruments, it can be seen that the breathing air that escapes from the bell can be detected over much shorter distances in comparison to the air that escapes during various activities (leaking air, intermittent exhalation, etc.), some of which are essential for sound production.

The range of the breathing air escaping from the bell of the brass instruments depends mainly on the width of the bore and the pressure of the breath with which the instrument is played. If dampers are used, the escaping airflow is restricted significantly. Exceptions are the French horn with a stopping mute and the F tuba with a damper, where the airflow is transported at least as far as when playing without a damper. Nevertheless, in most cases especially when using schlieren imaging, it can be seen that the escaping air either ascends due to natural convection or mixes with the surrounding room air and, therefore, cannot be visualized anymore.

The shape and range of the air escaping from the bell vary with regard to the specific physical characteristics of the musician and their blowing technique. Additionally, the angle at which the instrument is held determines the direction of the spreading air. Furthermore, it should be noted that the distances were measured from the bell of the instruments or the mouth of the singers (large font as well as French horn with stopping mute and F tuba with a damper, see Fig. 29) or from the mouth or mouthpiece from the musician (intermittent exhalation when playing the oboe or the bassoon, tone holes from the bassoon, air leak from the Bb clarinet, and air blown above the mouthpiece of the piccolo).

The results of the anemometry confirm the visualizations with both schlieren techniques. This is especially important as the schlieren technique can only detect flows based on a density gradient. The measured values do not fall below approximately 0.02 *m*/*s*. This is due to small movements of the surrounding air (e.g., due to the movements of fingers or hands when playing the instruments, air escaping from the tone holes, the musicians taking a breath between phrases, convective flows in the room, etc.). Furthermore, the measurements show that some of the measured velocities (e.g., oboe, grand flute, Bb trumpet, tenor trombone, and F tuba) decrease initially with increasing distance, but then increase again. This may be due to the turbulent properties of the flow which causes the velocity to vary due to the occurrence of small vortexes.

To reduce the breathing air escaping from the bell, especially of the brass instruments, a simple filter can also be used. In this case, the filter consists of cellulose and is taped directly to the bell of the instrument. This can be applied to every brass instrument because the breathing air fully escapes from the bell. For woodwind instruments, filters are not usable because the breathing air escapes not only from the bell but also from the tone holes and sometimes near the mouthpiece. As an outlook, Fig. 30b shows the spread of the breathing air when playing the trombone with the filter attached to the bell. Compared to Fig. 30a, where there is no barrier which could hamper the flow, the air spreads unhindered into the room.

**Fig. 30.**
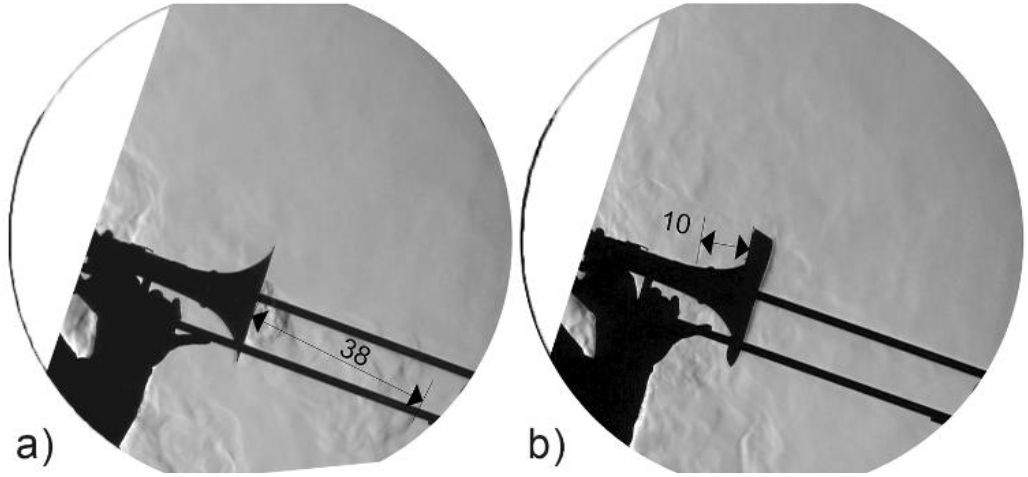
Trombone player with filter in front of the schlieren mirror (a), escaping breathing air while playing the tenor trombone without (b) and with filter (c)

Similar to the visualizations above, only the air escaping from the bell can be visualized. Thus, the actual spread of infectious particles cannot be estimated. However, it can be assumed that the filter works similar to speaking or singing while wearing a mask, where most of the particles stick to its material and, therefore, cannot enter the surrounding room air.

An exception to apply a filter to woodwind instruments are flutes. The visualizations in Fig. 14 showed that most of the breathing air is blown over the mouthpiece of the flutes. Hence, a filter can be placed in front of the mouthpiece to reduce the spread of the breathing air drastically (Fig. 31).

**Fig. 31.**
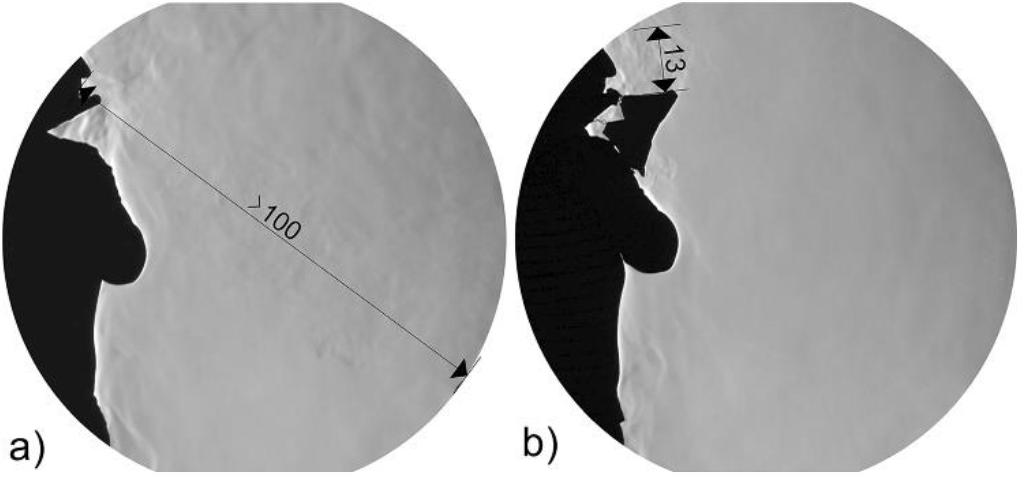
Range of the air blown over the mouthpiece of the grand flute without (a) and with filter (b)

## 5. Conclusion and limitations

The results presented in this article show the spread of the breathing air when playing wind instruments and singing. The results contribute to estimating the range of potentially infectious breathing air, which is essential during critical health periods such as the current COVID-19 pandemic. To visualize the escaping breathing air, the optical schlieren imaging system as well as the background-oriented schlieren (BOS) technique were used. However, these methods only visualize refractive index gradients that occur due to density gradients in transparent media. The schlieren imaging and BOS do not show the spread of small droplets or aerosols, which can contain pathogens. Therefore, the results can only be used to determine how far larger droplets could be transported with the exhaled air to define distance rules.

In the measurements shown above, the spread of the breathing air from only professional musicians and classical trained singers was investigated. However, musicians who are still learning how to play an instrument or how to sing may induce different spreading patterns of the exhaled air. As they may leak additional air near the mouthpiece of the played instrument or exhale more air than needed when singing, a larger amount of potentially infectious breathing air may spread into the room.

Another limitation of this study was the constant movement of the players during the velocity measurements even though they were asked to remain as still as possible. This affects the distance between the sensors and the bell or the mouth. Moreover, the movement of the instrument can increase the measured value. The velocity measurements showed that the breathing air escapes only at small velocities from the bell of the wind instrument. Here, differences can be observed within the diameter of the bell and the breathing pressure with which the instrument is played. For the air escaping from the mouth (exhaling when playing the oboe or bassoon and singing) and the air leaking near the mouthpiece (clarinet and bass clarinet), the air escapes at a higher velocity up to ν ≈ 0.15 *m*/*s*.

Using the visualizations and findings discussed in this article, the range, dimensions, and velocities of the escaping breathing air can be estimated. In addition to possible future investigation of the viral load of the escaping air, a holistic understanding of the possible risk posed by wind instruments and singers can be obtained.

## Data Availability

The data generated and analyzed during the current study are available on request from the corresponding author.
Supplementary videos can be found at the following links.

https://vimeo.com/431505952

https://vimeo.com/445131873

## 7. Acknowledgements

The authors would like to kindly acknowledge the commitment of the chief conductor and the wind players from the philharmonic orchestra *Thüringen Philharmonie Gotha – Eisenach* as well as the orchestra director, the wind players, and singers from the *Deutsches Nationaltheater und Staatskapelle Weimar*. We would also like to thank the German Research Foundation DFG for funding this project. Their support is highly appreciated.

